# Developing and validating polygenic risk scores for colorectal cancer risk prediction in East Asians

**DOI:** 10.1101/2022.02.14.22270878

**Authors:** Jie Ping, Yaohua Yang, Wanqing Wen, Sun-Seog Kweon, Koichi Matsuda, Wei-Hua Jia, Aesun Shin, Yu-Tang Gao, Keitaro Matsuo, Jeongseon Kim, Dong-Hyun Kim, Sun Ha Jee, Qiuyin Cai, Zhishan Chen, Ran Tao, Min-Ho Shin, Chizu Tanikawa, Zhi-Zhong Pan, Jae Hwan Oh, Isao Oze, Yoon-Ok Ahn, Keum Ji Jung, Zefang Ren, Xiao-Ou Shu, Jirong Long, Wei Zheng

## Abstract

**Importance:** Several polygenic risk scores (PRSs) have been developed to predict the risk of colorectal cancer (CRC); however, virtually all these PRSs were constructed in European descendants.

**Objective:** To develop and validate PRSs using data from large genome-wide association studies (GWAS) conducted in East Asians.

**Design, Setting, and Participants:** PRSs were developed using GWAS data from 22,702 cases and 212,486 controls and validated in two case-control studies (1,454 Korean and 1,736 Chinese). PRSs were derived using three approaches: (1) leading risk variants in GWAS-identified CRC loci; (2) risk variants independently associated with CRC in East Asians that were identified via fine-mapping of known GWAS-identified risk loci; and (3) genome-wide risk prediction algorisms using LDpred2 and PRS-CS.

**Main Outcomes and Measures:** Logistic regression models were used to examine associations of PRSs with CRC risk by estimating odds ratios (ORs) with 95% confidence intervals (CIs) and area under the receiver operating characteristic curve (AUC).

**Results:** In the validation sets, PRS_115-EAS_, a PRS with 115 GWAS-reported leading risk variants derived from East Asian data performed significantly better in discriminating CRC cases from non-cases than PRS_115-EUR_, a PRS derived using GWAS data from European descendants. In the Korea validation set, ORs per SD increase of PRS_115-EAS_ were 1.63 (95%CI = 1.46 - 1.82; AUC = 0.63), compared with OR of 1.44 (95%CI = 1.29 - 1.60, AUC = 0.60) for PRS_115-EUR_. PRS_115-EAS/EUR_ derived using results from meta-analyses of East Asian and European-ancestry data slightly improved the AUC to 0.64 (95% CI = 0.61 - 0.67). Similar, but less strong, associations were found in the China validation set. Individuals in the top 5% of PRS_115-EAS/EUR_ were at a 2.52-folded (95% CI = 2.27 - 2.81) elevated risk of CRC as compared with the average risk group and have a 12% or higher risk of developing CRC by age 85 years.

**Conclusions and Relevance:** Our results indicate that PRSs derived using GWAS-identified CRC risk variants are promising in predicting CRC risk in East Asians. Our study highlights the importance of using population-specific data to build CRC risk prediction models.

**Key Points:** *Question:* How well do polygenetic scores (PRS) developed using data from genome-wide association studies (GWAS) predict colorectal cancer (CRC) risk in East Asians?

*Findings:* Using GWAS data from more than 230,000 cases and controls in East Asian, 11 PRSs were developed with variants ranging from 115 to 747,643. PRS including 115 variants derived from meta-analyses of East Asian and European-ancestry GWAS data showed the highest performance in discriminating CRC cases and controls in validation sets including 1,490 cases and 1,700 controls.

*Meaning:* These findings support the utility of PRS in identifying high-risk individuals for CRC prevention and highlight the importance of using population-specific data to build CRC risk prediction models.

## Introduction

Colorectal cancer (CRC) is one of the most commonly diagnosed malignancies around the world.^1,2^ In the United States and many other developed countries, CRC incidence and mortality have steadily declined over the past few decades due to the implementation of effective population-based screening programs to detect and remove pre-cancerous lesions and early-stage cancer.^2-5^ However, the incidence and mortality of this cancer continue to rise in many Asian countries where rates are lower than those of the United States. Currently, many Asian countries do not have a population-based CRC screening program, significantly hindering the prevention of this disease. Because of economic constraints and differences in CRC risk, population-based CRC screening programs currently implemented in the U.S. and European countries may not be feasible or even appropriate for Asian countries. A cost-efficient, population-specific CRC screening strategy for Asians is imminently needed.

Current guidelines for initiating CRC screening are mainly based on age and family history of CRC, while more than 80% of CRC cases occur in individuals without a positive family history.^6^ CRC has a sizable heritable fraction and is a polygenic disease.^7^ Since 2007, genome-wide association studies (GWAS), including our studies conducted in East Asians^8-12^, identified common genetic variants in more than 190 loci associated with CRC risk.^8-24^ Polygenic risk scores (PRSs) constructed using CRC-associated risk variants as a measure of cumulative effect of these variants were evaluated for CRC risk prediction in several studies conducted in European-ancestry populations.^25-29^ However, to date, no study has systematically developed and validated PRSs for CRC risk prediction in East Asian populations. Given the difference in genetic architectures between Asian- and European-ancestry populations, PRSs established in European-ancestry populations may not perform well in predicting CRC risk in Asians. In 2009, we established the Asia Colorectal Cancer Consortium (ACCC) to evaluate genetic susceptibility factors of CRC in Asians. In the current study, we used GWAS data from the ACCC, including 24,192 CRC cases and 214,186 controls, to develop and validate performance of PRS in CRC risk prediction.

## Methods

### Datasets

To develop CRC risk prediction models for individuals of East Asian ancestry, we used GWAS data collected from 22,702 cases and 212,486 controls shown in Table 1. These study participants were recruited from eight studies conducted in China, Korea, and Japan. Data from the Korean-National Cancer Center CRC Study (Korea validation set: a case-control study including 622 cases and 832 controls) and a case-control study nested in cohorts of the Shanghai Men’s Health Study and Shanghai Women’s Health Study (China validation set: 868 cases and 868 age- and sex-frequency matched controls) were used for model validation. Detailed information on each study was previously reported^8-12^ and is described in the Supplementary Materials.

**Table 1.**
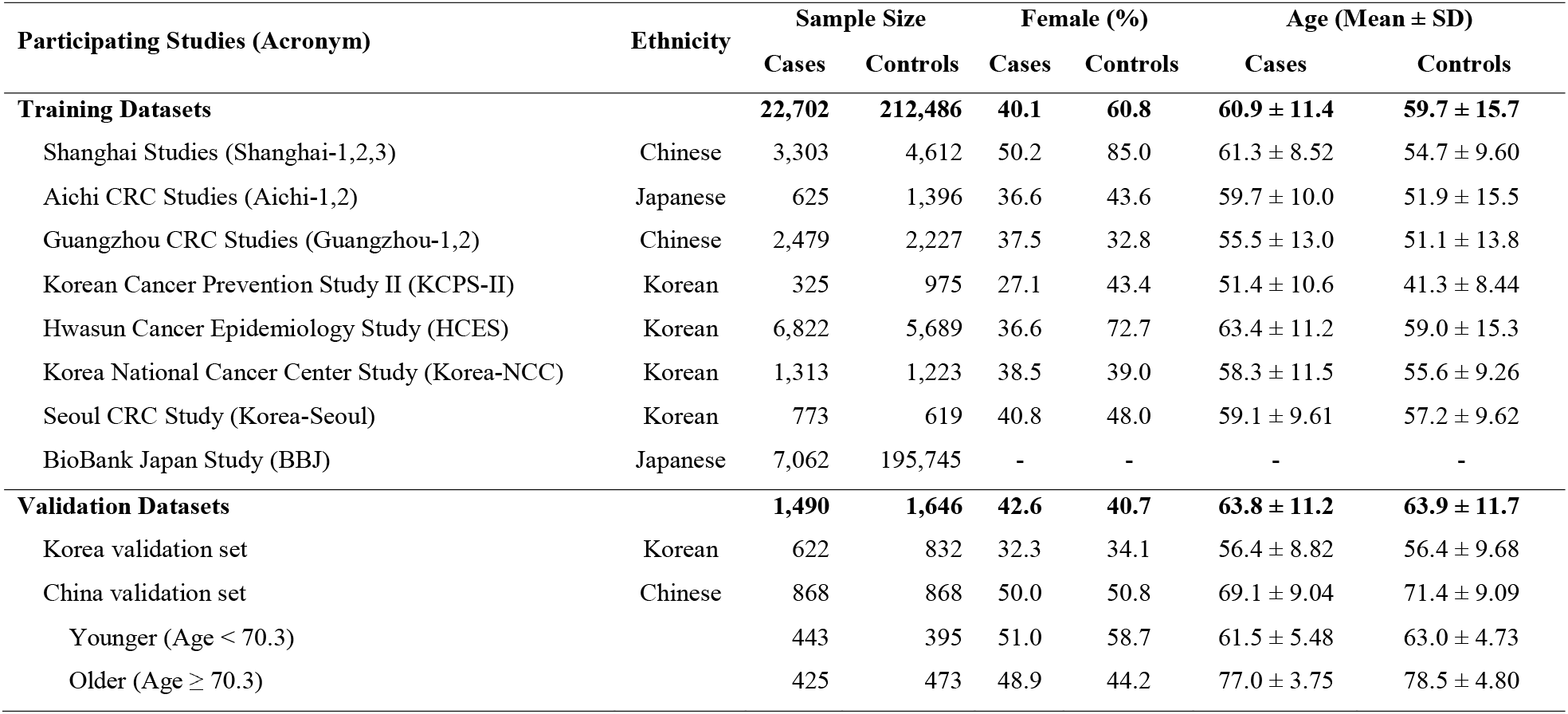
Sample size and selected descriptive statistics of participating studies: the Asia Colorectal Cancer Consortium (ACCC).

| Participating Studies (Acronym) | Ethnicity | Sample Size | | Female (%) | | Age (Mean ± SD) | |
| --- | --- | --- | --- | --- | --- | --- | --- |
|  |  | Cases | Controls | Cases | Controls | Cases | Controls |
| **Training Datasets** |  | **22,702** | **212,486** | **40.1** | **60.8** | **60.9 ± 11.4** | **59.7 ± 15.7** |
| Shanghai Studies (Shanghai-1,2,3) | Chinese | 3,303 | 4,612 | 50.2 | 85.0 | 61.3 ± 8.52 | 54.7 ± 9.60 |
| Aichi CRC Studies (Aichi-1,2) | Japanese | 625 | 1,396 | 36.6 | 43.6 | 59.7 ± 10.0 | 51.9 ± 15.5 |
| Guangzhou CRC Studies (Guangzhou-1,2) | Chinese | 2,479 | 2,227 | 37.5 | 32.8 | 55.5 ± 13.0 | 51.1 ± 13.8 |
| Korean Cancer Prevention Study II (KCPS-II) | Korean | 325 | 975 | 27.1 | 43.4 | 51.4 ± 10.6 | 41.3 ± 8.44 |
| Hwasun Cancer Epidemiology Study (HCES) | Korean | 6,822 | 5,689 | 36.6 | 72.7 | 63.4 ± 11.2 | 59.0 ± 15.3 |
| Korea National Cancer Center Study (Korea-NCC) | Korean | 1,313 | 1,223 | 38.5 | 39.0 | 58.3 ± 11.5 | 55.6 ± 9.26 |
| Seoul CRC Study (Korea-Seoul) | Korean | 773 | 619 | 40.8 | 48.0 | 59.1 ± 9.61 | 57.2 ± 9.62 |
| BioBank Japan Study (BBJ) | Japanese | 7,062 | 195,745 | - | - | - | - |
| **Validation Datasets** |  | **1,490** | **1,646** | **42.6** | **40.7** | **63.8 ± 11.2** | **63.9 ± 11.7** |
| Korea validation set | Korean | 622 | 832 | 32.3 | 34.1 | 56.4 ± 8.82 | 56.4 ± 9.68 |
| China validation set | Chinese | 868 | 868 | 50.0 | 50.8 | 69.1 ± 9.04 | 71.4 ± 9.09 |
| Younger (Age < 70.3) | | 443 | 395 | 51.0 | 58.7 | 61.5 ± 5.48 | 63.0 ± 4.73 |
| Older (Age ≥ 70.3) | | 425 | 473 | 48.9 | 44.2 | 77.0 ± 3.75 | 78.5 ± 4.80 |

### Genotyping and Imputation

Details of genotyping, quality control, and imputation for the ACCC were reported previously^8-12^ and are provided in the Supplementary Materials (Supplementary Table S1). We imputed genotype data using the 1000 Genomes Project Phase III data as a reference via the Michigan Imputation Server.^30^ Only variants with minor allele frequency (MAF) > 5%, a high imputation quality (*R*^*2*^ > 0.8), and presented in over half of our training datasets were included for further analyses.

### Polygenic Risk Score Calculation

To estimate odds ratios of SNPs associated with CRC risk, we used logistic regression models adjusting for age, sex, and top 10 principal components (PCs). Association analyses were performed in each of the eight ACCC datasets included in the training set, and then a meta-analysis was conducted to estimate pooled odds ratios using METAL software.^31^ PRSs were calculated as weighted sums of alleles associated with CRC risk using equation 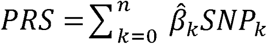 where SNP_k_ was the allelic dosage and 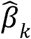 was the corresponding log-odds ratio of SNP_k_ associated with CRC risk derived from the meta-analysis. We developed PRSs using three different approaches as described below and summarized in Figure 1.

**Figure 1.**
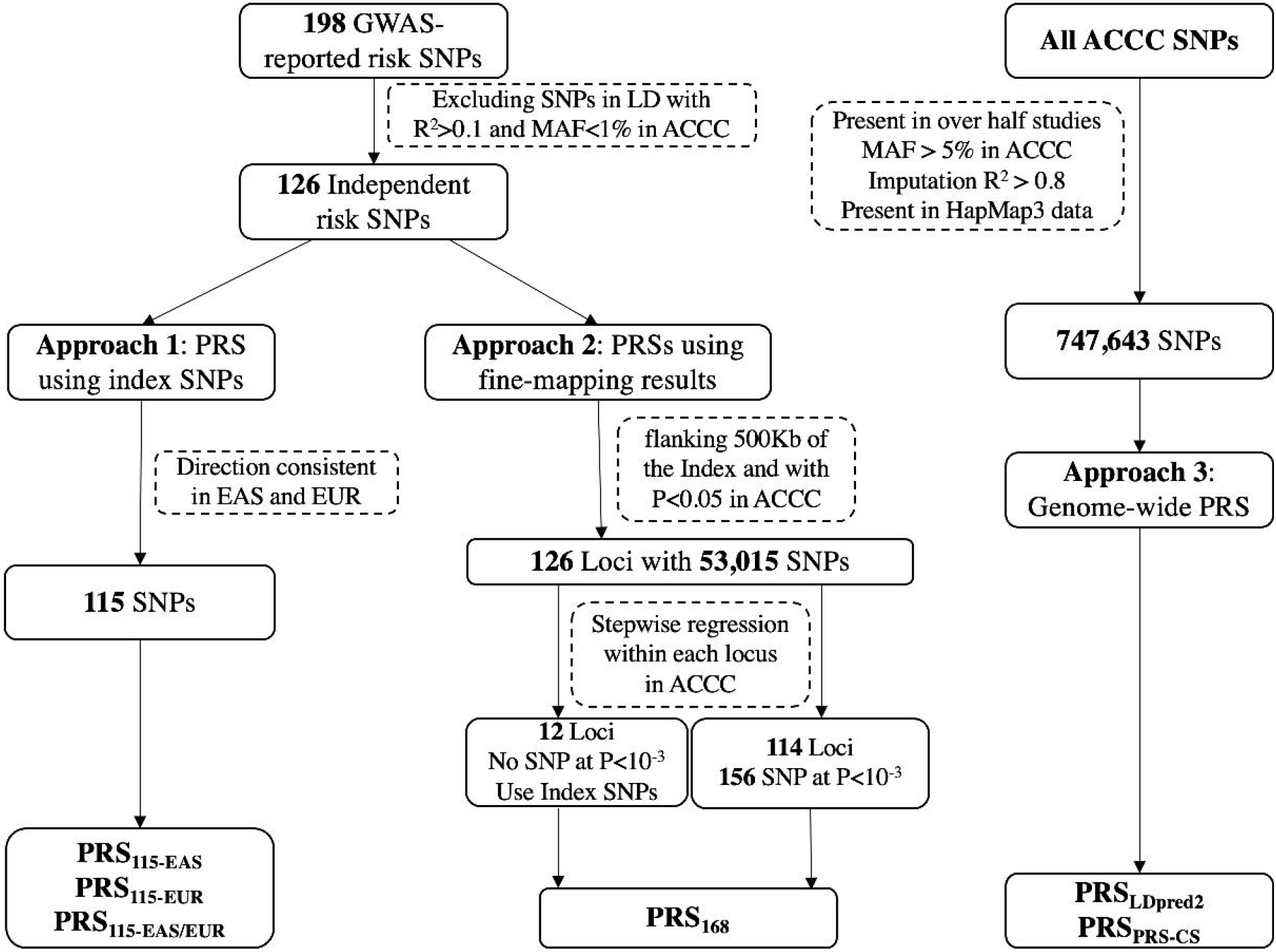
Summary of approaches to derive polygenic risk scores (PRS) for colorectal cancer (CRC) in East Asians.

#### Approach 1: PRS using GWAS-identified index risk SNPs

By searching the literature, we identified common SNPs in 198 regions showing a significant association with CRC risk at *P* < 5.0 × 10^−8^ from large CRC GWASs conducted in East Asians and European-ancestry populations.^8-24^ If two SNPs are in linkage disequilibrium (LD) with *R*^*2*^ > 0.1 in East Asians (1000 Genomes, Phase 3 v5), the SNP with lower P value was kept for PRS construction. In total, 126 SNPs with MAF > 1% were included as independent CRC risk variants. Of them, 115 risk variants were consistent in the direction of association with CRC risk in both East Asian and European descendants, and these risk variants were used to construct PRS_115_. European log-odds ratios obtained from the literature were used as the weights to construct PRS_115-EUR_, while log-odds ratios derived from our ACCC training set were used to construct PRS_115-EAS_. We performed a meta-analysis of log-odds ratios derived from European and East Asian populations using the fixed-effects model to estimate the pooled log-odds ratios, and used them as weights to construct PRS_115-EAS/EUR_.

#### Approach 2: PRS using SNPs selected from fine-mapping of GWAS-identified risk loci

Approximately 78% (n=99) of these 126 independent CRC risk variants were initially identified in GWAS conducted in European descendants. To identify additional independent risk variants and risk variants more strongly associated with CRC in each of these regions in East Asians, we performed fine-mapping analyses with data from our training set using the Genome-wide Complex Trait Analysis (GCTA) method described by Yang.^32^ We first extracted 53,015 SNPs located at flanking 500Kbp regions of each of the 126 index SNPs and with nominal significant association at *P* < 0.05 in the training set. We then conducted conditional and joint analyses (COJO) in each region adjusting for the corresponding GWAS-identified lead SNP, from which we identified 156 SNPs of 114 loci showing an independent association with CRC risk at *P* < 1.0 × 10^−3^. Adjusted log-odds ratios of these 156 SNPs, along with log-odds ratios of the GWAS-identified risk variants for the remaining 12 loci where no SNP showed an association with CRC risk at *P* < 1.0 × 10^−3^ in our training data set, were used to construct the PRS (PRS_168_). We also evaluated PRSs conducted using risk SNPs selected using more stringent p-values (such as 1.0 × 10^−4^ and 1.0 × 10^−5^), and results for these evaluations are shown in Supplementary Table S3.

#### Approach 3: PRSs based on genome-wide risk prediction algorithms

We used LDpred and PRS-CS to derive genome-wide PRSs using data from the training set. LDpred is a Bayesian approach that considers LD among SNPs^33^ and may have a higher accuracy than PRS using only GWAS-identified risk SNPs.^27^ We used summary statistics from our ACCC training set for model training. LD matrix was calculated using genotype data of all ACCC training sets except the BioBank Japan study (BBJ), which included 32,381 samples. According to LDpred2 recommendation, we restricted our analysis to SNPs included in the HapMap3 SNPs. After applying these criteria, 747,643 SNPs were included in this analysis. Log-odds ratios of these SNPs with CRC risk were re-estimated using LDpred2 with default settings. We applied four models from LDpred2 in this study (infinitesimal model <LDpred2-Inf>, the best model among all sparse models <LDpred2-grid-sp>, the best model among all non-sparse models <LDpred2-grid-nosp>, and the automatic model <LDpred2-Auto>).

PRS-CS is a Bayesian approach that infers posterior SNP effect sizes under continuous shrinkage (CS) priors using summary statistics and an external LD reference panel.^34^ Summary statistics from our ACCC training set and pre-calculated LD reference panel of East Asians constructed using the UK Biobank data from PRS-CS were used for model training.

### Model Performance Assessment and Risk Estimation

We evaluated the performance of PRSs derived in training sets in our validation datasets by calculating the area under the receiver operating characteristic curve (AUC). Odds ratio (OR) and 95% confidence interval (CI) per one standard deviation (SD) increase in PRS were estimated using logistic regression. Age, sex, ethnic group, study sites, and top 10 PCs were adjusted in the risk estimation model using logistic regression. We also estimated ORs for selected PRS groups relative to those in the average risk group (40^th^ to 60^th^ PRS percentile). We estimated ten-year and lifetime absolute risk of CRC by PRS categories for Chinese, Korean, and Japanese subjects using ORs from our study of the association of PRS with CRC risk and data on CRC incidence and mortality rates for China, Japan, and South Korea obtained from the GLOBOCAN database (2020)^1^. All statistical analyses were conducted using R v4.1.2.

## Results

Data from eight datasets were used for PRS training and two datasets for PRS validation. Selected characteristics of study participants are summarized by studies in Table 1. Cases and controls differed considerably by age and sex in multiple studies included in the training set and thus these two variables were adjusted for in the analysis. In the validation sets, age and sex were comparable between cases and controls.

As described in the method section, we developed PRS_115-EAS,_ PRS_115-EUR,_ and PRS_115-EAS/EUR_ using log-odds ratios for 115 GWAS-identified risk variants derived from GWAS conducted in East Asians, European-ancestry populations, and meta-analyses of both populations, respectively. These log-odds ratios were used as weights for constructing PRS_115_ (Supplementary Table S2). PRS_115-EAS_ performed significantly better in discriminating CRC cases from non-cases than PRS_115-EUR_. In the Korea validation set, ORs per SD increase of PRS_115-EAS_ were 1.63 (95%CI = 1.46 - 1.82; AUC = 0.63), compared with OR of 1.44 (95%CI = 1.29 - 1.60, AUC = 0.60) for PRS_115-EUR_. PRS_115-EAS/EUR_ slightly improved the AUC to 0.64 (95% CI = 0.61 - 0.67). Similar but weaker associations were found in the China validation set. Because subjects included in the China validation set were older than those included in the Korea validation set, we performed stratified analyses by age using the mean age (70.3 years) as the cut-off to evaluate if the performance of PRSs differ by age. The performance of all three PRSs was better in the younger group than the older group in the China validation set (Supplementary Table S3). For example, the PRS_115-EAS/EUR_ showed an AUC of 0.63 (95% CI = 0.60 - 0.67) and OR of 1.62 (1.40 – 1.88) in the younger China validation set, while its AUC in the older group is only 0.59 (0.56 - 0.63) and the OR is 1.39 (1.21 - 1.60). The difference between these two OR estimates was statistically significant (P for interaction, 0.038).

Using fine-mapping methods, we developed three PRSs based on COJO-p-values cut-off of 1.0 × 10^−3^, 1.0 × 10^−4^, and 1.0 × 10^−5^ to select SNPs for PRS construction (Supplementary Table S3). Of the three PRSs evaluated, PRS_168_, based on p-value of 1.0 × 10^−3^, had the best performance, showing an AUC of 0.62 and 0.58 in the Korea and China validation sets, respectively (Table 2). Performances of other two PRSs in the validation sets were similar. None of these PRSs performed better than the PRS_115-EAS_ or PRS_115-EAS/EUR_ described above.

**Table 2.** Associations of polygenic risk scores with colorectal cancer risk in the validation datasets.

| PRS development methods | Korea validation set |  | China validation set |  |
| --- | --- | --- | --- | --- |
|  | OR (95% CI)<sup>1</sup> | AUC (95% CI) | OR (95% CI)<sup>1</sup> | AUC (95% CI) |
| **GWAS-reported index SNPs<sup>2</sup>** |  |  |  |  |
| **PRS<sub>115-EAS</sub>** | 1.63 (1.46 - 1.83) | 0.63 (0.60 - 0.66) | 1.51 (1.37 - 1.67) | 0.61 (0.59 - 0.64) |
| **PRS<sub>115-EUR</sub>** | 1.44 (1.29 - 1.60) | 0.60 (0.57 - 0.63) | 1.39 (1.26 - 1.53) | 0.59 (0.56 - 0.61) |
| **PRS<sub>115-EAS/EUR</sub>** | 1.68 (1.50 - 1.89) | 0.64 (0.61 - 0.67) | 1.50 (1.36 - 1.66) | 0.61 (0.58 - 0.64) |
| **SNPs selected by fine-mapping<sup>3</sup>** |  |  |  |  |
| **PRS<sub>168</sub>** | 1.54 (1.38 - 1.72) | 0.62 (0.59 - 0.65) | 1.32 (1.20 - 1.45) | 0.58 (0.55 - 0.61) |
| **Genome-wide risk prediction algorithm<sup>4</sup>** |  |  |  |  |
| **PRS<sub>LDpred2-Grid-Sp</sub>** | 1.43 (1.29 - 1.60) | 0.60 (0.57 - 0.63) | 1.32 (1.20 - 1.45) | 0.58 (0.55 - 0.60) |
| **PRS<sub>PRS-CS</sub>** | 1.34 (1.20 - 1.49) | 0.58 (0.55 - 0.61) | 1.35 (1.23 - 1.49) | 0.58 (0.55 - 0.61) |

\* PRS, polygenic risk score; OR, odds ratio; CI, confidence interval; AUC, area under the receiver operating characteristic curve.
1. Odds ratio and 95% confidence interval per standard deviation increase in PRS were estimated using logistic regression. 2. All weights for European ancestry (PRS<sub>115-EUR</sub>) were obtained from published papers. Weights for East Asian (PRS<sub>115-EAS</sub>) were derived from the ACCC training set. Weights of PRS<sub>115-EAS/EUR</sub> were from a meta-analysis based on weights for European ancestry and weights for East Asians. 3. Weights for PRS were derived from the ACCC training set. 4. Weights for PRS based on LDpred2 and PRS-CS were re-estimated using LDpred2 and PRS-CS based on the ACCC training set.

We developed five PRSs using the genome-wide risk prediction algorithms, based on four LDpred2 models (infinitesimal model, the best model among all sparse models, the best model among all non-sparse models, and the automatic model) and one PRS-CS model to derive weights for SNPs for PRS construction. Of these five PRSs evaluated, PRS_LDpred2-Grid-Sp_, based on the best main model among all sparse models, demonstrated the highest discriminative ability, showing an AUC of 0.60 and 0.58 in the Korea and China validation sets, respectively (Table 2). Results based on PRS-CS (PRS_PRS-CS_) showed AUCs of 0.58 in both the Korea and China validation sets. Other PRS performances in the validation sets were similar and are shown in Supplementary Table S3. Again, none of these PRSs performed better than the PRS_115-EAS_ or PRS_115-EAS/EUR_ described above.

Because the PRS_115-EAS/EUR_ showed the best performance in our external validation, subsequent analyses were based on this PRS. We estimated ORs according to percentile of PRS_115-EAS/EUR_ in Table 3. A clear dose-response association between PRS levels and CRC risk was observed in the combined validation dataset, including samples from both Korea and China (*P for trend < 0*.*001*). Because of a relatively small sample size in the validation set, we also evaluated the associations of PRS level with CRC risk in all subjects included in the ACCC (including both training and validation datasets, except BBJ data). The risk estimates were similar in these two datasets, except those at the 95^th^ to 99^th^ and >99^th^ percentile groups, in which the estimated ORs were higher in the combined dataset than the validation set. This difference is likely due to unstable estimates in the validation set because of a small sample size.

**Table 3.** Odds ratios of colorectal cancer risk in association with PRS_115-EAS/EUR_ in the validation datasets^1^ and ACCC datasets combined^2^.

| PRS Percentile | Validation datasets |  | ACCC datasets combined |  |
| --- | --- | --- | --- | --- |
|  | Number of Cases/Controls | OR (95% CI) | Number of Cases/Controls | OR (95% CI) |
| ≤1 | 5/18 | 0.33 (0.11 - 0.85) | 35/177 | 0.34 (0.23 - 0.49) |
| (1, 5] | 21/68 | 0.37 (0.22 - 0.61) | 183/683 | 0.48 (0.40 - 0.57) |
| (5, 10] | 34/85 | 0.51 (0.33 - 0.78) | 269/858 | 0.56 (0.48 - 0.65) |
| (10, 20] | 80/170 | 0.58 (0.42 - 0.79) | 639/1713 | 0.66 (0.59 - 0.73) |
| (20, 40] | 226/339 | 0.81 (0.64 - 1.02) | 1573/3426 | 0.82 (0.75 - 0.89) |
| (40, 60] | 278/340 | 1.00 (Reference) | 1930/3425 | 1.00 (Reference) |
| (60, 80] | 332/339 | 1.19 (0.96 - 1.49) | 2577/3426 | 1.34 (1.24 - 1.45) |
| (80, 90] | 223/170 | 1.59 (1.23 - 2.05) | 1494/1712 | 1.56 (1.43 - 1.71) |
| (90, 95] | 128/85 | 1.93 (1.41 - 2.66) | 917/857 | 1.88 (1.68 - 2.10) |
| (95, 99] | 122/68 | 2.21 (1.58 - 3.11) | 915/683 | 2.34 (2.09 - 2.63) |
| >99 | 34/18 | 2.29 (1.28 - 4.24) | 332/177 | 3.22 (2.65 - 3.92) |
| **P for trend** |  | < 0.001 |  | < 0.001 |
| **Top 5% Vs Remaining** |  | 2.21 (1.68 - 2.92) |  | 2.40 (2.19 - 2.64) |
| **Top 1% Vs Remaining** |  | 2.17 (1.23 - 3.95) |  | 2.92 (2.42 - 3.53) |

1. Including 1,490 cases and 1,646 controls from both Korea and China validation sets.
2. Including 1,7130 cases and 18,387 controls from both training and validation datasets except BBJ data. Individual genotype data for BBJ were not available for the present study.

We used incidence and mortality data from the GLOBOCAN database for the year 2020 to estimate absolute CRC risks by PRS group. Figure 2 shows the estimated 10-year absolute risk of CRC by PRS_115-EAS/EUR_ groups in Chinese, Japanese, and Korean subjects. The recommended age to start colorectal cancer screening in the general population is 40 years in Japan ^35^ and 50 years in both China ^36^ and South Korea ^37^. At age 50, the 10-year absolute risks for CRC were 0.47% and 0.52% for average-risk individuals (at 40-60^th^ PRS percentile) in China and Korea, respectively (Supplementary Table S5). In Japan, the 10-year absolute risk for CRC was 0.24% for an average risk individual at age 40. However, individuals in the top 1% PRS_115-EAS/EUR_ group (>99^th^ percentile) reached this risk level by ages 38, 36, and 32 in China, Korea, and Japan, respectively, much earlier than the average risk group.

**Figure 2.**
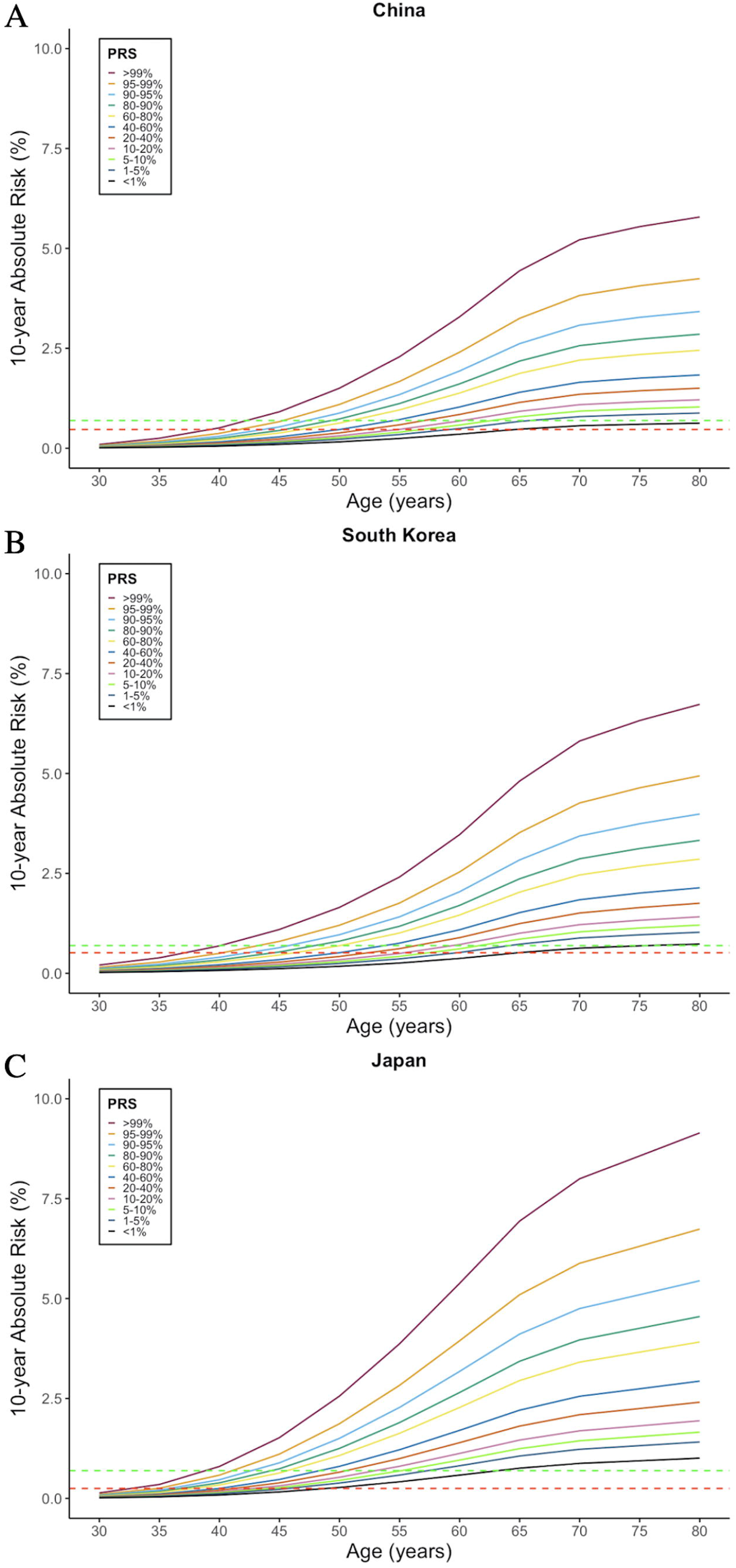
Ten-year absolute risks of colorectal cancer (CRC) by PRS_115-EAS/EUR_ groups in China, South Korea, and Japan. The green horizontal lines show the 10-year absolute CRC risk (0.69%) for individuals at age 50 in the United States in the year 2020. The red horizontal lines show the 10-year absolute CRC risk for individuals at age 50 in China (0.47%, Figure 2A) and South Korea (0.52% Figure 2B), and 10-year absolute CRC risk (0.24%, Figure 2C) for individuals at age 40 in Japan.

The 10-year absolute CRC risk is 0.69% for average-risk individuals at age 50 in the United States. An individual with a medium PRS_115-EAS/EUR_ (40^th^ to 60^th^ percentile) reaches this risk level at age 54, 53, and 48 years in China, Korea, and Japan, respectively (Figure 2). However, among those in the top 1% risk group, an individual would reach this risk level at age 42, 40, and 38 years in China, Korea, and Japan, respectively.

The estimated lifetime absolute risks for East Asians by PRS_115-EAS/EUR_ categories for CRC are shown in Figure 3. By age 85, the absolute risk of CRC in the top 1% of PRS_115-EAS/EUR_ was 16.4%, 18.6%, and 26.0%, in China, Korea, and Japan, respectively. In the lowest 1% of PRS_115-EAS/EUR_, by age 85, the absolute risk of CRC was 1.7%, 2.0%, and 2.7% in China, Korea, and Japan, respectively. Individuals in the top 5% of PRS_115-EAS/EUR_ were estimated to have 12.9%, 14.5%, and 20.4% risk of developing CRC by age 85 years in China, Korea, and Japan, respectively.

**Figure 3.**
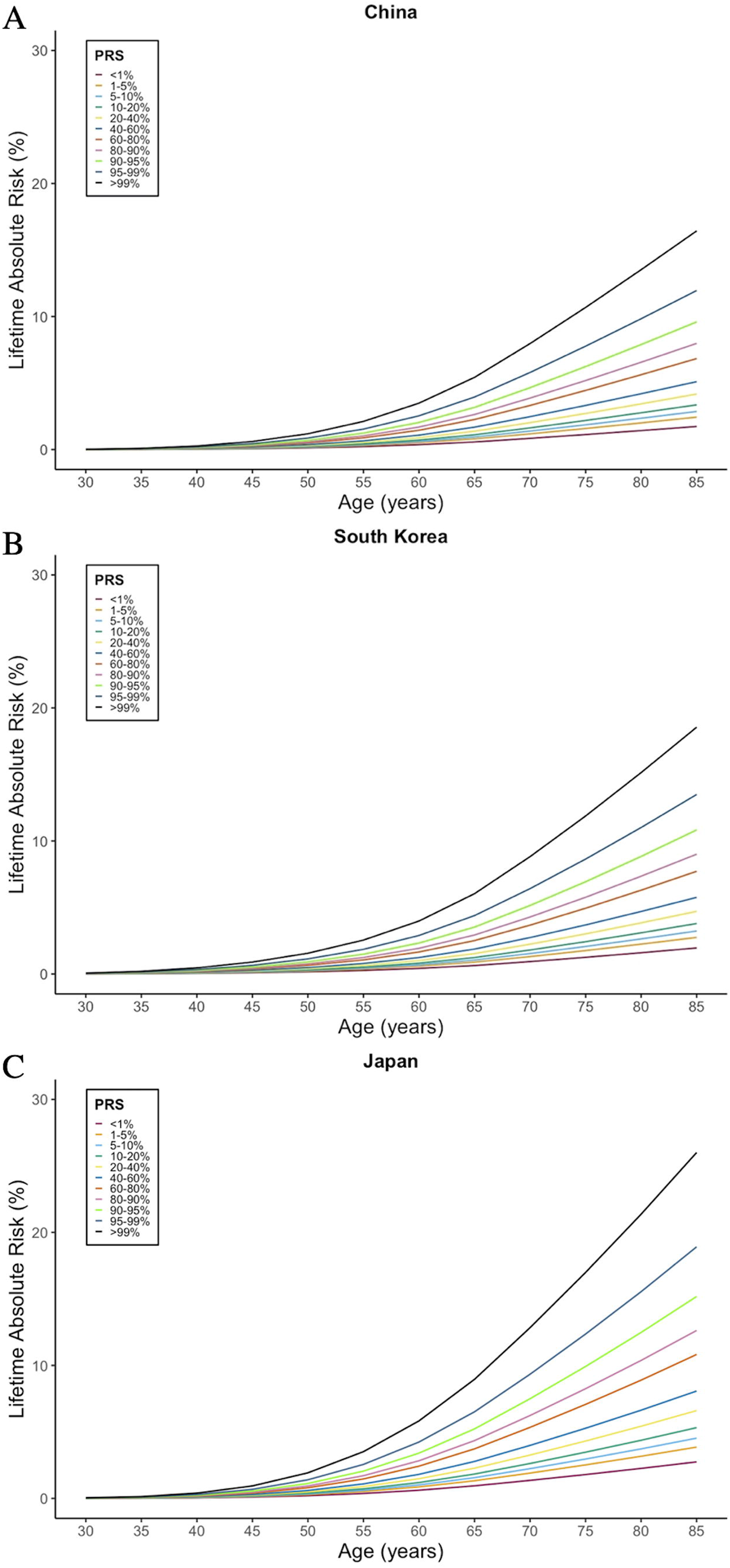
Lifetime absolute risks of colorectal cancer by PRS_115-EAS/EUR_ groups in China, South Korea, and Japan.

## Discussion

In this study, we built and validated PRSs to predict CRC risk for East Asians using the largest data set available to date, including 24,192 CRC cases and 214,186 controls of East Asian ancestry from the ACCC. We found that the PRSs derived using GWAS-identified CRC risk variants showed promise in predicting the risk of this common cancer, particularly when data from East Asians were used in constructing the PRS. Our study demonstrates the importance of incorporating population-specific data to build risk prediction models for CRC.

Theoretically, one would expect that PRSs built using risk variants selected from fine-mapping known CRC risk loci or using genome-wide algorithms would outperform PRS constructed using only GWAS-reported CRC risk variants. In our study, however, these PRSs showed a poorer performance than PRSs derived using GWAS-reported risk variants. The sample sizes used in fine-mapping and deriving genome-wide PRSs were relatively small, which might have affected the stability of risk estimates in this study. Therefore, future studies with a larger sample size are needed to further improve the performance of PRS in East Asians.

An interesting finding from our study is that the discriminative ability of PRS was lower in older study participants. For example, the AUC for PRS_115-EAS/EUR_ was 0.64 in the Korea validation set (mean age = 56.4). In the China validation set (mean age = 70.3), however, the PRS_115-EAS/EUR_ had a poor performance with an AUC of 0.61. When stratified by age in the China validation set, we showed that the performance of PRS_115-EAS/EUR_ is better in the younger group (AUC = 0.63, mean age = 62.2) than the older group (AUC = 0.60, mean age = 77.8). Our findings for a significant interaction between age and PRS were supported by recent studies conducted in European-ancestry populations.^27,38^ It is possible that lifestyle and environmental risk factors may play a more significant role in the etiology of CRC in older than younger patients, and thus the prediction accuracy of PRS may decrease among elderly compared with younger populations.

Although this is the largest study ever conducted to date in East Asians to develop and validate CRC risk prediction using GWAS data for this cancer, the sample size is relatively small, particularly in analyses conducted by ethnic groups. Because of the small sample size for the validation set, we used log-odds ratios derived from all data combined to estimate absolute risk. This could lead to potential overfitting. However, the ORs of CRC by PRS percentile estimated using data from the validation set or all ACCC data were similar, suggesting that overfitting may not be a major concern.

In summary, we found that PRSs derived using GWAS data of CRC showed promise in predicting the risk of this cancer in East Asians, particularly when data from East Asians are included in constructing the PRS. Our study demonstrated the need to use population-specific data to build risk prediction models in CRC. The predictive accuracy of PRS developed in our study remains moderate and could be improved in future studies with a larger sample size.

## Data Availability

All data produced in the present study are available upon reasonable request to the authors

## Acknowledgements

The authors thank all study participants and research staff of all parent studies for their contributions and commitment to this project. The authors Ms. Rachel Mullen for editing the manuscript. The work at Vanderbilt University Medical Center was supported by U.S. NIH grants R01CA188214, R37CA070867, UM1CA182910, R01CA124558, R01CA158473, and R01CA148667, as well as Anne Potter Wilson Chair funds from the Vanderbilt University School of Medicine. Sample preparation and genotyping assays at Vanderbilt University were conducted at the Survey and Biospecimen Shared Resources and Vanderbilt Microarray Shared Resource, supported in part by the Vanderbilt-Ingram Cancer Center (P30CA068485). Statistical analyses were performed on servers maintained by the Advanced Computing Center for Research and Education (ACCRE) at Vanderbilt University (Nashville, TN). Studies (listed with grant support) participating in the Asia Colorectal Cancer Consortium include the Shanghai Women’s Health Study (US NIH, R37CA070867, UM1CA182910), the Shanghai Men’s Health Study (US NIH, R01CA082729, UM1CA173640), the Shanghai Breast and Endometrial Cancer Studies (US NIH, R01CA064277 and R01CA092585; contributing only controls), the Shanghai Colorectal Cancer Study 3 (US NIH, R37CA070867, R01CA188214 and Anne Potter Wilson Chair funds), the Guangzhou Colorectal Cancer Study (National Key Scientific and Technological Project, 2011ZX09307-001-04; the National Basic Research Program, 2011CB504303, contributing only controls, the Natural Science Foundation of China, 81072383, contributing only controls), the Hwasun Cancer Epidemiology Study–Colon and Rectum Cancer (HCES-CRC; grants from Chonnam National University Hwasun Hospital Biomedical Research Institute, HCRI18007), the Japan BioBank Colorectal Cancer Study (grant from the Ministry of Education, Culture, Sports, Science and Technology of the Japanese government), the Aichi Colorectal Cancer Study (Grant-in-Aid for Cancer Research, grant for the Third Term Comprehensive Control Research for Cancer and Grants-in-Aid for Scientific Research from the Japanese Ministry of Education, Culture, Sports, Science and Technology, 17015018 and 221S0001), the Korea-NCC (National Cancer Center) Colorectal Cancer Study (Basic Science Research Program through the National Research Foundation of Korea, 2010-0010276 and 2013R1A1A2A10008260; National Cancer Center Korea, 0910220), and the KCPS-II Colorectal Cancer Study (National R&D Program for Cancer Control, 1631020; Seoul R&D Program, 10526).

## Supplementary Tables

**Table S1**. Genotyping platforms of participating studies: Asia Colorectal Cancer Consortium (ACCC).

**Table S2**. Log odds ratios for GWAS-reported risk SNPs from the literature (EUR), ACCC (EAS), and meta-analysis (EAS/EUR).

**Table S3**. Associations of all polygenic risk scores with colorectal cancer risk in the validation sets and the age-stratified China validation set.

**Table S4**. 10-Year Absolute Risk (%) for people at age 50 within different PRS_115-EAS/EUR_ groups in China, South Korea, and Japan.

## References

1. Sung H, Ferlay J, Siegel RL, et al. Global Cancer Statistics 2020: GLOBOCAN Estimates of Incidence and Mortality Worldwide for 36 Cancers in 185 Countries. CA Cancer J Clin. May 2021;71(3):209–249. doi:10.3322/caac.21660

2. Howlader N, Noone A, Krapcho M, et al. SEER Cancer Statistics Review, 1975-2018. National Cancer Institute Bethesda, MD. 2021;

3. Sandouk F, Al Jerf F, Al-Halabi MH. Precancerous lesions in colorectal cancer. Gastroenterol Res Pract. 2013;2013:457901. doi:10.1155/2013/457901

4. Rawla P, Sunkara T, Barsouk A. Epidemiology of colorectal cancer: incidence, mortality, survival, and risk factors. Prz Gastroenterol. 2019;14(2):89–103. doi:10.5114/pg.2018.81072

5. Wong MC, Ding H, Wang J, Chan PS, Huang J. Prevalence and risk factors of colorectal cancer in Asia. Intest Res. Jul 2019;17(3):317–329. doi:10.5217/ir.2019.00021

6. Smith RA, Cokkinides V, von Eschenbach AC, et al. American Cancer Society guidelines for the early detection of cancer. CA Cancer J Clin. Jan-Feb 2002;52(1):8–22. doi:10.3322/canjclin.52.1.8

7. Lichtenstein P, Holm NV, Verkasalo PK, et al. Environmental and heritable factors in the causation of cancer--analyses of cohorts of twins from Sweden, Denmark, and Finland. N Engl J Med. Jul 13 2000;343(2):78–85. doi:10.1056/NEJM200007133430201

8. Jia WH, Zhang B, Matsuo K, et al. Genome-wide association analyses in East Asians identify new susceptibility loci for colorectal cancer. Nat Genet. Feb 2013;45(2):191–6. doi:10.1038/ng.2505

9. Zhang B, Jia WH, Matsuda K, et al. Large-scale genetic study in East Asians identifies six new loci associated with colorectal cancer risk. Nat Genet. Jun 2014;46(6):533–42. doi:10.1038/ng.2985

10. Zeng C, Matsuda K, Jia WH, et al. Identification of Susceptibility Loci and Genes for Colorectal Cancer Risk. Gastroenterology. Jun 2016;150(7):1633–1645. doi:10.1053/j.gastro.2016.02.076

11. Lu Y, Kweon SS, Tanikawa C, et al. Large-Scale Genome-Wide Association Study of East Asians Identifies Loci Associated With Risk for Colorectal Cancer. Gastroenterology. Apr 2019;156(5):1455–1466. doi:10.1053/j.gastro.2018.11.066

12. Lu Y, Kweon SS, Cai Q, et al. Identification of Novel Loci and New Risk Variant in Known Loci for Colorectal Cancer Risk in East Asians. Cancer Epidemiol Biomarkers Prev. Feb 2020;29(2):477–486. doi:10.1158/1055-9965.EPI-19-0755

13. Dunlop MG, Dobbins SE, Farrington SM, et al. Common variation near CDKN1A, POLD3 and SHROOM2 influences colorectal cancer risk. Nat Genet. May 27 2012;44(7):770–6. doi:10.1038/ng.2293

14. Houlston RS, Cheadle J, Dobbins SE, et al. Meta-analysis of three genome-wide association studies identifies susceptibility loci for colorectal cancer at 1q41, 3q26.2, 12q13.13 and 20q13.33. Nat Genet. Nov 2010;42(11):973–7. doi:10.1038/ng.670

15. Huyghe JR, Bien SA, Harrison TA, et al. Discovery of common and rare genetic risk variants for colorectal cancer. Nat Genet. Jan 2019;51(1):76–87. doi:10.1038/s41588-018-0286-6

16. Law PJ, Timofeeva M, Fernandez-Rozadilla C, et al. Association analyses identify 31 new risk loci for colorectal cancer susceptibility. Nat Commun. May 14 2019;10(1):2154. doi:10.1038/s41467-019-09775-w

17. Orlando G, Law PJ, Palin K, et al. Variation at 2q35 (PNKD and TMBIM1) influences colorectal cancer risk and identifies a pleiotropic effect with inflammatory bowel disease. Hum Mol Genet. Jun 1 2016;25(11):2349–2359. doi:10.1093/hmg/ddw087

18. Schmit SL, Edlund CK, Schumacher FR, et al. Novel Common Genetic Susceptibility Loci for Colorectal Cancer. J Natl Cancer Inst. Feb 1 2019;111(2):146–157. doi:10.1093/jnci/djy099

19. Schmit SL, Schumacher FR, Edlund CK, et al. A novel colorectal cancer risk locus at 4q32.2 identified from an international genome-wide association study. Carcinogenesis. Nov 2014;35(11):2512–9. doi:10.1093/carcin/bgu148

20. Study C, Houlston RS, Webb E, et al. Meta-analysis of genome-wide association data identifies four new susceptibility loci for colorectal cancer. Nat Genet. Dec 2008;40(12):1426–35. doi:10.1038/ng.262

21. Tenesa A, Farrington SM, Prendergast JG, et al. Genome-wide association scan identifies a colorectal cancer susceptibility locus on 11q23 and replicates risk loci at 8q24 and 18q21. Nat Genet. May 2008;40(5):631–7. doi:10.1038/ng.133

22. Wang H, Burnett T, Kono S, et al. Trans-ethnic genome-wide association study of colorectal cancer identifies a new susceptibility locus in VTI1A. Nat Commun. Aug 8 2014;5:4613. doi:10.1038/ncomms5613

23. Wang M, Gu D, Du M, et al. Common genetic variation in ETV6 is associated with colorectal cancer susceptibility. Nat Commun. May 5 2016;7:11478. doi:10.1038/ncomms11478

24. Whiffin N, Hosking FJ, Farrington SM, et al. Identification of susceptibility loci for colorectal cancer in a genome-wide meta-analysis. Hum Mol Genet. Sep 1 2014;23(17):4729–37. doi:10.1093/hmg/ddu177

25. Smith T, Gunter MJ, Tzoulaki I, Muller DC. The added value of genetic information in colorectal cancer risk prediction models: development and evaluation in the UK Biobank prospective cohort study. Br J Cancer. Oct 2018;119(8):1036–1039. doi:10.1038/s41416-018-0282-8

26. Dunlop MG, Tenesa A, Farrington SM, et al. Cumulative impact of common genetic variants and other risk factors on colorectal cancer risk in 42,103 individuals. Gut. Jun 2013;62(6):871–81. doi:10.1136/gutjnl-2011-300537

27. Thomas M, Sakoda LC, Hoffmeister M, et al. Genome-wide Modeling of Polygenic Risk Score in Colorectal Cancer Risk. Am J Hum Genet. Sep 3 2020;107(3):432–444. doi:10.1016/j.ajhg.2020.07.006

28. Khera AV, Chaffin M, Aragam KG, et al. Genome-wide polygenic scores for common diseases identify individuals with risk equivalent to monogenic mutations. Nat Genet. Sep 2018;50(9):1219–1224. doi:10.1038/s41588-018-0183-z

29. Hsu L, Jeon J, Brenner H, et al. A model to determine colorectal cancer risk using common genetic susceptibility loci. Gastroenterology. Jun 2015;148(7):1330–9 e14. doi:10.1053/j.gastro.2015.02.010

30. Das S, Forer L, Schonherr S, et al. Next-generation genotype imputation service and methods. Nat Genet. Oct 2016;48(10):1284–1287. doi:10.1038/ng.3656

31. Willer CJ, Li Y, Abecasis GR. METAL: fast and efficient meta-analysis of genomewide association scans. Bioinformatics. Sep 1 2010;26(17):2190–1. doi:10.1093/bioinformatics/btq340

32. Yang J, Lee SH, Goddard ME, Visscher PM. GCTA: a tool for genome-wide complex trait analysis. Am J Hum Genet. Jan 7 2011;88(1):76–82. doi:10.1016/j.ajhg.2010.11.011

33. Vilhjalmsson BJ, Yang J, Finucane HK, et al. Modeling Linkage Disequilibrium Increases Accuracy of Polygenic Risk Scores. Am J Hum Genet. Oct 1 2015;97(4):576–92. doi:10.1016/j.ajhg.2015.09.001

34. Ge T, Chen CY, Ni Y, Feng YA, Smoller JW. Polygenic prediction via Bayesian regression and continuous shrinkage priors. Nat Commun. Apr 16 2019;10(1):1776. doi:10.1038/s41467-019-09718-5

35. Hamashima C. Cancer screening guidelines and policy making: 15 years of experience in cancer screening guideline development in Japan. Jpn J Clin Oncol. Mar 1 2018;48(3):278–286. doi:10.1093/jjco/hyx190

36. Fang JY, Zheng S, Jiang B, et al. Consensus on the Prevention, Screening, Early Diagnosis and Treatment of Colorectal Tumors in China: Chinese Society of Gastroenterology, October 14-15, 2011, Shanghai, China. Gastrointest Tumors. Jun 2014;1(2):53–75. doi:10.1159/000362585

37. Luu XQ, Lee K, Lee YY, Suh M, Kim Y, Choi KS. Acceptance on colorectal cancer screening upper age limit in South Korea. World J Gastroenterol. Jul 21 2020;26(27):3963–3974. doi:10.3748/wjg.v26.i27.3963

38. Archambault AN, Su YR, Jeon J, et al. Cumulative Burden of Colorectal Cancer-Associated Genetic Variants Is More Strongly Associated With Early-Onset vs Late-Onset Cancer. Gastroenterology. Apr 2020;158(5):1274–1286 e12. doi:10.1053/j.gastro.2019.12.012

