## Supplementary Materials and Tables for "Developing and validating polygenic risk scores for colorectal cancer risk prediction in East Asians"

**eMethods.**

**eReferences.**

**eTable1.**

**eTable2.**

**eTable3.**

**eTable4.**

**Developing and validating polygenic risk scores for colorectal cancer risk prediction in East Asians**

Jie Ping^1^, Yaohua Yang^1^, Wanqing Wen^1^, Sun-Seog Kweon^2^, Koichi Matsuda^3^, Wei-Hua Jia^4^, Aesun Shin^5, 6^, Yu-Tang Gao^7^, Keitaro Matsuo^8, 9^, Jeongseon Kim^10^, Dong-Hyun Kim^11^, Sun Ha Jee^12^, Qiuyin Cai^1^, Zhishan Chen^1^, Ran Tao^13^, Min-Ho Shin^2^, Chizu Tanikawa^14^, Zhi-Zhong Pan^4^, Jae Hwan Oh^15^, Isao Oze^16^, Yoon-Ok Ahn^17^, Keum Ji Jung^12^, Zefang Ren^18^, Xiao-Ou Shu^1^, Jirong Long^1^, Wei Zheng^1, *^

^1^ Division of Epidemiology, Department of Medicine, Vanderbilt-Ingram Cancer Center, Vanderbilt Epidemiology Center, Vanderbilt University School of Medicine, Nashville, TN, USA;
^2^ Department of Preventive Medicine, Chonnam National University Medical School, Gwangju, South Korea;
^3^ Laboratory of Clinical Genome Sequencing, Department of Computational Biology and Medical Sciences, Graduate School of Frontier Sciences, University of Tokyo, Tokyo, Japan;
^4^ State Key Laboratory of Oncology in South China, Cancer Center, Sun Yat-sen University, Guangzhou, China;
^5^ Department of Preventive Medicine, Seoul National University College of Medicine, Seoul, Korea;
^6^ Cancer Research Institute, Seoul National University, Seoul, Korea;
^7^ State Key Laboratory of Oncogenes and Related Genes and Department of Epidemiology, Shanghai Cancer Institute, Renji Hospital, Shanghai Jiaotong University School of Medicine, Shanghai, China;
^8^ Division of Molecular and Clinical Epidemiology, Aichi Cancer Center Research Institute, Nagoya, Japan;
^9^ Department of Epidemiology, Nagoya University Graduate School of Medicine, Nagoya, Japan;
^10^ Department of Cancer Biomedical Science, Graduate School of Cancer Science and Policy, National Cancer Center, Gyeonggi-do, South Korea;
^11^ Department of Social and Preventive Medicine, Hallym University College of Medicine, Okcheon-dong, Korea;
^12^ Department of Epidemiology and Health Promotion, Graduate School of Public Health, Yonsei University, Seoul, Korea;
^13^ Department of Biostatistics, Vanderbilt University, 37212 Nashville, TN, USA;
^14^ Laboratory of Genome Technology, Human Genome Center, Institute of Medical Science, University of Tokyo, Tokyo, Japan;
^15^ Center for Colorectal Cancer, National Cancer Center Hospital, National Cancer Center, Gyeonggi-do, South Korea;
^16^ Division of Cancer Epidemiology and Prevention, Aichi Cancer Center Research Institute, Nagoya, Japan;
^17^ Department of Preventive Medicine, Seoul National University College of Medicine, Seoul, South Korea;
^18^ School of Public Health, Sun Yat-sen University, Guangzhou, China

*** Corresponding author**:

**Wei Zheng**, M.D., Ph.D.
Vanderbilt Epidemiology Center

Vanderbilt University School of Medicine

2525 West End Avenue, 8^th^ Floor, Nashville, TN 37203-1738


**eMethods**

**Study Participants**

This study included 24,192 CRC cases and 214,186 controls from the Asia Colorectal Cancer Consortium (ACCC) that were recruited from eight centers located in China, South Korea, and Japan. The protocols for all participating studies were approved by the relevant review boards at their respective institutions. Provided below are brief descriptions for each study.

**The Shanghai Colorectal Cancer Study:** This study included 4,171 cases and 7,071 controls from four case-control sets conducted in Shanghai, China (Shanghai-1, Shanghai-2, Shanghai-3 and Shanghai-4). CRC cases for the Shanghai-1, Shanghai-2, and Shanghai-4 studies were identified from two cohort studies: the Shanghai Women’s Health Study (SWHS, n = 921) and the Shanghai Men’s Health Study (SMHS, n = 663). Cancer-free controls were identified from the SWHS (n = 4,928) and the SMHS (n = 857) [Shanghai-1, Shanghai-2 and Shanghai-4]. The SWHS and the SMHS are both population-based cohort studies conducted in urban Shanghai, China. The SWHS includes 75,049 Chinese women between the ages of 40 and 70 years at enrollment (1997 to 2000) and who lived in urban Shanghai. In-person interviews were conducted to collect exposure information, and anthropometrics were measured. The response rate was 92% for the baseline interview. Approximately 88% of the study participants provided biological samples, either a blood sample (n = 56,833) or an exfoliated buccal cell sample (n = 8,921). Using similar study protocols, the SMHS enrolled 61,582 men between the ages of 40 and 74 years in urban Shanghai between 2001 and 2006, with an overall response rate of 74% for the baseline interview. Approximately 90% of the study participants provided either a blood sample (76%) or a buccal cell sample (14%). Ongoing follow-up for cancer incidence and cause-specific mortality is being conducted for both the SMHS and SWHS via a combination of periodic in-person surveys and annual linkage with data routinely collected by the population-based Shanghai Cancer Registry and Vital Statistics Unit (for death certificates). CRC cases for the Shanghai-3 study were identified from the population-based Shanghai Cancer Registry. In-person interviews and saliva sample collections were completed for 2,587 cases. Controls (n = 1,286) for these cases were selected from members of the SWHS and SMHS who were free of any cancer.

**The Guangzhou Colorectal Cancer Study:** This study included 2,479 cases and 2,227 controls from two case-control studies conducted in Guangzhou, China (Guangzhou-1 and Guangzhou-2). Cases were recruited from the Sun Yat-Sen University Cancer Center, Guangdong, China, between January 2002 and January 2012. Controls were cancer-free men and women recruited from the physical examination centers of several large hospitals in Guangdong during the same time period.

**The Aichi Colorectal Cancer study:** This study included 625 cases and 1,396 controls from two case-control studies conducted in Aichi, Japan (n = 1,340 in Aichi-1 and n = 681 in Aichi-2). It was conducted as part of the Hospital-based Epidemiologic Research Program at the Aichi Cancer Center in Japan. All study participants (n = 28,766) were recruited between December 2000 and November 2005. CRC case status was confirmed via the Hospital-based Epidemiologic Research Program database, as well as the hospital-based cancer registry at the Aichi Cancer Center^1^.

**BioBank Japan:** This study (BBJ) includes 7,062 cases and 195,745 controls. DNA samples of both CRC cases and cancer-free controls were obtained from the BioBank Japan of the Personalized Medicine Project (<http://biobankjp.org/>).

**The Korean Cancer Prevention Study-II:** This study **(**KCPS-II**)** included 325 cases and 975 controls. It was conducted as part of the Korean Cancer Prevention Study-II, a cohort study with 266,258 individuals, aged 20-77, who visited 16 health promotion centers nationwide between April 2004 and December 2008 in South Korea^2^. Cancer status was confirmed via the National Cancer Registry and hospitalization records. Cancer-free controls were randomly selected from the same cohort study. Details of this study have been described elsewhere^2,3^.

**The Hwasun Cancer Epidemiology Study - Colon and Rectum Cancer (HCES-CRC):** HCES-CRC consisted of two studies, the first of which included 3,130 cases and 4,625 controls, and the second of which (HCES2-CRC) included 3,465 cases and 1,445 controls. These were hospital-based case-control studies conducted in South Korea that included multiple cancers^4,5^. Cases were CRC patients at Chonnam National University Hwasun Hospital, Jeollanam-do, South Korea, newly diagnosed between April 2004 and October 2014. Cancer-free controls were randomly selected from participants in the Korean Community Health Survey, an annual nationwide health interview survey, conducted from 2010 to 2012 in the Jindo and Bosung counties, Jeollanam-do, South Korea^6^.

**The Korean-National Cancer Center Colorectal Cancer Study:** This study had two collections (Korea-NCC and Korea-NCC2) with a total of 1,935 cases and 2,055 controls. Korea-NCC included 1,313 cases and 1,223 controls; Korea-NCC2 included 622 cases and 832 controls. It was a hospital-based case-control study of CRC, conducted in South Korea. For Korea-NCC, cases were histologically confirmed patients with CRC who received surgery between 2000 and 2004 at the Korean National Cancer Center (NCC). A total of 1,935 patients participated and provided a blood sample. Controls (n = 2,055) were selected from participants of the Cancer Screening Cohort of the NCC, recruited between August 2002 and December 2004. For Korea-NCC2, cases included newly diagnosed colorectal cancer patients at the Center for Colorectal Cancer of the National Cancer Center, Korea, between August 2010 and August 2013. The control participants were recruited between October 2007 and December 2014 among individuals visiting the Center for Cancer Prevention and Detection at the same hospital for a health check-up program provided by the National Health Insurance Cooperation, which covers the entire Korean population.

**The Seoul Colorectal Cancer Study:** This study (Korea-Seoul) is a multicenter case-control study conducted in South Korea^7^. Cases were CRC patients who were admitted to two university hospitals or one general cancer hospital in the Seoul Metropolitan Area between 1995 and 2004^7^. Controls were patients of the same hospitals during the same time period from a wide spectrum of inpatients with non-neoplastic conditions. A total of 773 cases and 619 controls providing a blood sample were included in Stage 2 of this project.

**Laboratory Procedures**

Methods of genotyping, genotype calling, and quality control for Shanghai-1, Shanghai-2, Guangzhou-1, KCPS-II, and Aichi-1 have been described elsewhere^2,3,8-17^. Briefly, genotyping was performed using the Affymetrix Genome-Wide Human SNP Array 6.0 (Affymetrix Inc., Santa Clara, CA, United States) for Shanghai-1 cases and controls; the Illumina Human OmniExpress BeadChip (Illumina Inc., San Diego, CA, United States) for Shanghai-2 cases and controls, Guangzhou-1 cases and Aichi-1 cases; the Illumina Infinium HumanHap550 BeadChip (Illumina Inc., San Diego, CA, United States) and the Illumina 660W-Quad BeadChip (Illumina Inc., San Diego, CA, United States) for Shanghai-2 controls; the Illumina Human610-Quad BeadChip (Illumina Inc., San Diego, CA, United States) for Guangzhou-1 controls; the Illumina Infinium HumanHap610 BeadChip (Illumina Inc., San Diego, CA, United States) for Aichi-1 controls and the Affymetrix Genome-Wide Human SNP Array 5.0 (Affymetrix Inc., Santa Clara, CA, United States) for KCPS-II cases and controls. Genotype calling was performed using the Birdseed algorithm v2 for Affymetrix arrays or GenomeStudio software for Illumina arrays, based on the manufacturers’ protocols. We only included shared SNPs in the analysis if samples in a study were genotyped on different platforms. As described previously, a uniform quality control protocol was employed to exclude samples and SNPs from each of the Stage 1 studies^3,9,10,18^, and samples or SNPs were excluded if they met any of the following criteria: (i) genotype call rate per sample < 95%, (ii) genetically identical or duplicate samples (i.e., PI_HAT > 0.9), (iii) sex determined using genotypes inconsistent with epidemiological or clinical data, (iv) first- or second-degree relatives (i.e., PI_HAT > 0.25), (v) ethnic outliers with a population structure inconsistent with HapMap Asian samples (see Evaluation of population structure and Supplementary Figure 1)^15^, (vi) genotype call rate per SNP < 95%, (vii) minor allele frequency (MAF) < 5%, (viii) genotyping consistency rates < 95% in quality control samples, (ix) *P* for Hardy-Weinberg equilibrium (HWE) <1×10^-5^ in controls, or (x) SNPs not in autosomes. After these quality control filtering procedures, 500,547 SNPs for 3,100 individuals (474 cases and 2,626 controls) remained in the Shanghai-1 dataset; 245,386 SNPs for 904 individuals (254 cases and 650 controls) remained in the Shanghai-2 dataset; 250,011 SNPs for 1,609 participants (638 cases and 971 controls) remained in the Guangzhou-1 dataset; 232,426 SNPs for 1,340 participants (401 cases and 939 controls) remained in the Aichi-1 dataset; and 312,869 SNPs for 1,300 participants (325 cases and 975 controls) remained in the KCPS-II dataset.

For Guangzhou-2, genotyping was performed using Illumina HumanExome-12v1 Beadchip (Illumina Inc., San Diego, CA, United States), either at the Genome Quebec Innovation Centre (Montreal, Quebec, Canada) or at the Shanghai Genergy Biotechnology, following Illumina’s protocol. Genotype calling was carried out using Illumina’s GenTrain version 2.0 clustering algorithm in GenomeStudio. A total of 33,285 samples plus 192 QC samples were genotyped to create the cluster files, and over 35,000 samples were called together. Cluster boundaries were determined using study samples. After clustering, approximately 80,000 variants were manually reviewed and clusters were edited for 27,506 variants. We evaluated concordance rates for HapMap samples genotyped in our study and sequenced by the 1000 Genomes Project. Principal components analyses were conducted based on 3,200 ancestry informative markers on the Exomechip using EIGENSTRAT to identify population outliers with the 1000 Genomes Project data as a reference^15^. We also estimated the pairwise proportion of identify-by-descent to identify duplicated samples or close relatives. The samples were excluded if (i) the call rate < 98%, (ii) they were heterozygosity outliers, (iii) ethnic outliers, (iv) samples with close relationships, (v) their consistence rates among duplicated samples < 99%, or (vi) the samples were with the wrong sex. The SNPs were excluded if the (i) MAF < 0.1%, (ii) call rate < 98%, (iii) genotyping concordance rate < 98%, (iv) HWE test *P* < 10^-5^, or (v) cautious SNPs were discovered by the Exomechip design group. The final dataset included 71,534 variants on 1,841 CRC cases and 1,256 controls for Guangzhou-2.

Details on genotyping and quality control procedures for BBJ have been previously reported for the Japanese GWAS,^9,10,15,18,19^ and some procedures are updated herein. Briefly, both cases and controls in BBJ were genotyped using the Illumina HumanHap610-Quad BeadChip or OmniExpressExome (Illumina Inc., San Diego, CA, United States). Samples were excluded according to the following criteria: (i) genotype call rate of < 99%, (ii) genetically identical or duplicated samples, (iii) sex determined using genetic data inconsistent with epidemiological or clinical data, (iv) first- or second-degree relatives, (v) ancestry outliers, or (vi) heterozygosity outliers. SNPs were excluded using the following criteria: (i) genotype call rate of < 99%, (ii) MAF < 1%, (iii) difference in allelic frequency between Japan Biobank and 1000 Genomes > 0.16, or (iv) SNPs with *P* for HWE < 1 × 10^−6^ in controls. We obtained the final datasets, including 512,115 SNPs genotyped in 6,692 cases and 27,178 controls. Genotyping for Shanghai-3 and HCES-CRC was completed using either Illumina Infinium HTS iSelect Consortium-OncoArray^20^ in Shanghai Genergy Biotechnology (Shanghai-3) or the University of Southern California Molecular Genomics Core Facility (HCES-CRC), following Illumina’s protocol. Raw data were imported into GenomeStudio and genotypes were called using cluster definitions provided by the OncoArray Consortium. Samples were excluded according to the following criteria: (i) genotype call rate of < 95%, (ii) genetically identical or duplicated samples, (iii) sex determined using genetic data inconsistent with epidemiological or clinical data, (iv) first- or second-degree relatives, (v) ancestry outliers, or (vi) heterozygosity outliers. SNPs were excluded using the following criteria: (i) genotype call rate of < 95%, (ii) one of duplicate probes with the low call rate, (iii) SNPs with non-ideal cluster plots, (iv) SNPs with *P* for HWE < 1 × 10^−7^ in controls or with *P* for HWE < 1 × 10^−12^ in cases. We obtained the final datasets, including 475,512 SNPs genotyped in 2,575 cases and 1,336 controls from the Shanghai CRC Study 3, and 474,697 SNPs genotyped in 3,130 cases and 4,625 controls from the HCES-CRC study.

Cases and controls for Shanghai-4, Aichi-2, Korea-NCC, Korea-NCC2, Korea-Seoul, and HCES2-CRC were genotyped using the Illumina MEGA-Expanded Array (Illumina Inc., San Diego, CA, United States). Raw data were imported into GenomeStudio and genotypes were called using cluster definitions provided by Illumina. Samples were excluded according to the following criteria: (i) genotype call rate of < 95%, (ii) genetically identical or duplicated samples, (iii) sex determined using genetic data inconsistent with epidemiological or clinical data, (iv) first- or second-degree relatives, or (v) ancestry outliers. SNPs were excluded using the following criteria: (i) genotype call rate of < 95%, (ii) monomorphic SNPs, (iii) one of duplicate probes with the low call rate, (iv) SNPs with *P* for HWE < 1 × 10^−7^ in controls or with *P* for HWE < 1 × 10^−12^ in cases. After these quality control filtering procedures, 1,948,772 SNPs for 3,359 individuals (865 cases and 2,494 controls) remained in the Shanghai-4 dataset; 1,937,421 SNPs for 681 individuals (224 cases and 457 controls) remained in the Aichi-2 dataset; 1,937,421 SNPs for 2,536 participants (1,313 cases and 1,223 controls) remained in the Korea-NCC dataset; 1,937,421 SNPs for 1,454 participants (622 cases and 832 controls) remained in the Korea-NCC2 dataset; 1,937,421 SNPs for 1,392 participants (773 cases and 619 controls) remained in the Korea-Seoul dataset; and 1,944,529 SNPs for 5,964 participants (3,445 cases and 2,519 controls) remained in the HCES2-CRC dataset. Before the imputation, variants were further excluded for (i) MAF < 1%; (ii) difference in allelic frequency between study samples and 1000 Genome phase3 East Asian population is > 0.2, or (iii) SNPs not included in the 1000 Genome phase 3 project.

**Imputation**

To increase the coverage of genotypes, the respective quality-controlled samples from 13 studies (Shanghai-1, Shanghai-2, Shanghai-3, Shangha-4, Guangzhou-1, Aichi-1, Aichi-2, KCPS-II, HCES-CRC, Korea-NCC, Korea-NCC2, Korea-Seoul, and HCES2-CRC) were imputed using 1000 Genome phase 3 mixed reference haplotypes via the Michigan Imputation Server (SHAPIT2 for haplotype phasing and minimac3 for imputation). Dosage data were generated from imputation to represent the expected number of copies of the effect allele (a value between 0 and 2) for each SNP. For each study, we obtained approximately 47 million genotyped or imputed autosomal SNPs on the 22 autosomes. Imputation of the ungenotyped SNPs in BBJ samples was conducted with phased data of JPT/CHS/CHD subjects from the 1000 Genomes Project phase1 v3 (16-March-2012 release) as the reference using MACH v1.0 (<http://www.sph.umich.edu/csg/abecasis/MACH/>) and minimac (<http://genome.sph.umich.edu/wiki/Minimac>). Before imputation was performed, we removed SNPs from the reference panel if they met any of the following criteria: (i) MAF < 1%, (ii) P for HWE <10^-5^, or (iii) large allele frequency difference between BBJ and the reference panel (i.e., 0.16). After imputation, we obtained approximately 7.5 million genotyped or imputed autosomal SNPs.

**eReferences**

1.	Matsuo K, Suzuki T, Ito H, et al. Association between an 8q24 locus and the risk of colorectal cancer in Japanese. *BMC Cancer*. Oct 26 2009;9:379. doi:10.1186/1471-2407-9-379

2.	Jee SH, Sull JW, Lee JE, et al. Adiponectin concentrations: a genome-wide association study. *Am J Hum Genet*. Oct 8 2010;87(4):545-52. doi:10.1016/j.ajhg.2010.09.004

3.	Jia WH, Zhang B, Matsuo K, et al. Genome-wide association analyses in East Asians identify new susceptibility loci for colorectal cancer. *Nat Genet*. Feb 2013;45(2):191-6. doi:10.1038/ng.2505

4.	Song HR, Shin MH, Kim HN, et al. Sex-specific differences in the association between ABO genotype and gastric cancer risk in a Korean population. *Gastric Cancer*. Apr 2013;16(2):254-60. doi:10.1007/s10120-012-0176-z

5.	Cui LH, Shin MH, Kweon SS, et al. Methylenetetrahydrofolate reductase C677T polymorphism in patients with gastric and colorectal cancer in a Korean population. *BMC Cancer*. May 26 2010;10:236. doi:10.1186/1471-2407-10-236

6.	Ahn HR, Cho SB, Chung IJ, Kweon SS. Socioeconomic differences in self- and family awareness of viral hepatitis status among carriers of hepatitis B or C in rural Korea. *Am J Infect Control*. Mar 2018;46(3):328-332. doi:10.1016/j.ajic.2017.09.001

7.	Kim J, Cho YA, Kim DH, et al. Dietary intake of folate and alcohol, MTHFR C677T polymorphism, and colorectal cancer risk in Korea. *Am J Clin Nutr*. Feb 2012;95(2):405-12. doi:10.3945/ajcn.111.020255

8.	Zheng W, Long J, Gao YT, et al. Genome-wide association study identifies a new breast cancer susceptibility locus at 6q25.1. Research Support, N.I.H., Extramural

Research Support, U.S. Gov't, Non-P.H.S. *Nature genetics*. Mar 2009;41(3):324-8. doi:10.1038/ng.318

9.	Zhang B, Jia WH, Matsuda K, et al. Large-scale genetic study in East Asians identifies six new loci associated with colorectal cancer risk. *Nat Genet*. Jun 2014;46(6):533-42. doi:10.1038/ng.2985

10.	Zeng C, Matsuda K, Jia WH, et al. Identification of Susceptibility Loci and Genes for Colorectal Cancer Risk. *Gastroenterology*. Jun 2016;150(7):1633-45. doi:10.1053/j.gastro.2016.02.076

11.	Abnet CC, Freedman ND, Hu N, et al. A shared susceptibility locus in PLCE1 at 10q23 for gastric adenocarcinoma and esophageal squamous cell carcinoma. Research Support, N.I.H., Extramural

Research Support, N.I.H., Intramural. *Nature genetics*. Sep 2010;42(9):764-7. doi:10.1038/ng.649

12.	Bei JX, Li Y, Jia WH, et al. A genome-wide association study of nasopharyngeal carcinoma identifies three new susceptibility loci. Research Support, Non-U.S. Gov't. *Nature genetics*. Jul 2010;42(7):599-603. doi:10.1038/ng.601

13.	Nakata I, Yamashiro K, Yamada R, et al. Association between the SERPING1 gene and age-related macular degeneration and polypoidal choroidal vasculopathy in Japanese. Research Support, Non-U.S. Gov't. *PloS one*. Apr 19 2011;6(4):e19108. doi:10.1371/journal.pone.0019108

14.	Amundadottir L, Kraft P, Stolzenberg-Solomon RZ, et al. Genome-wide association study identifies variants in the ABO locus associated with susceptibility to pancreatic cancer. Letter. *Nature genetics*. Sep 2009;41(9):986-90. doi:10.1038/ng.429

15.	Lu Y, Kweon SS, Tanikawa C, et al. Large-scale Genome-wide Association Study of East Asians Identifies Loci Associated With Risk for Colorectal Cancer. *Gastroenterology*. Dec 6 2018;doi:10.1053/j.gastro.2018.11.066

16.	Lu Y, Kweon SS, Cai Q, et al. Identification of Novel Loci and New Risk Variant in Known Loci for Colorectal Cancer Risk in East Asians. *Cancer Epidemiol Biomarkers Prev*. Feb 2020;29(2):477-486. doi:10.1158/1055-9965.EPI-19-0755

17.	Lu Y, Kweon SS, Tanikawa C, et al. Large-Scale Genome-Wide Association Study of East Asians Identifies Loci Associated With Risk for Colorectal Cancer. *Gastroenterology*. Apr 2019;156(5):1455-1466. doi:10.1053/j.gastro.2018.11.066

18.	Zhang B, Jia WH, Matsuo K, et al. Genome-wide association study identifies a new SMAD7 risk variant associated with colorectal cancer risk in East Asians. Research Support, N.I.H., Extramural

Research Support, Non-U.S. Gov't. *International journal of cancer Journal international du cancer*. Aug 15 2014;135(4):948-55. doi:10.1002/ijc.28733

19.	Cui R, Okada Y, Jang SG, et al. Common variant in 6q26-q27 is associated with distal colon cancer in an Asian population. Meta-Analysis

Research Support, Non-U.S. Gov't. *Gut*. Jun 2011;60(6):799-805. doi:10.1136/gut.2010.215947

20.	Amos CI, Dennis J, Wang Z, et al. The OncoArray Consortium: A Network for Understanding the Genetic Architecture of Common Cancers. *Cancer epidemiology, biomarkers & prevention : a publication of the American Association for Cancer Research, cosponsored by the American Society of Preventive Oncology*. Jan 2017;26(1):126-135. doi:10.1158/1055-9965.EPI-16-0106

**eTables**

**eTable 1.** Genotyping platforms of participating studies: Asia Colorectal Cancer Consortium (ACCC).

| **Study** | **Genotyping platform** |
| --- | --- |
| **Training datasets for deriving PRS** | |
| Shanghai Colorectal Cancer Study | |
| Shanghai-1 | Affymetrix GenomeWide Human SNP Array 6.0 |
| Shanghai-2 | Illumina OmniExpress BeadChip |
| Shanghai-3 | Illumina Infinium OncoArray-500K BeadChip |
| BioBank Japan | |
|  | Illumina OmniExpress BeadChip |
| Aichi Colorectal Cancer Study | |
| Aichi-1 | Illumina OmniExpress BeadChip |
| Aichi-2 | Illumina Multi-Ethnic Genotyping Array |
| Guangzhou Colorectal Cancer Study | |
| Guangzhou-1 | Illumina OmniExpress BeadChip |
| Guangzhou-2 | Illumina HumanExome-12v1_A Beadchip |
| Korean Cancer Prevention Study-II (KCPS-II) | |
|  | Affymetrix GenomeWide Human SNP Array 5.0 |
| Hwasun Cancer Epidemiology Study - Colon and Rectum Cancer | |
| HCES-CRC | Illumina Infinium OncoArray-500K BeadChip |
| HCES2-CRC | Illumina Multi-Ethnic Genotyping Array |
| Korean-National Cancer Center Colorectal Cancer Study | |
| Korea-NCC | Illumina Multi-Ethnic Genotyping Array |
| Seoul Colorectal Cancer Study |  |
|  | Illumina Multi-Ethnic Genotyping Array |
| **Validation datasets for evaluating PRS performance** | |
| Korea validation set | Illumina Multi-Ethnic Genotyping Array |
| China validation set | Illumina Multi-Ethnic Genotyping Array |

**eTable 2.** Summary of results for GWAS-reported risk SNPs from the literature (EUR), ACCC (EAS), and meta-analysis (EAS/EUR).

| **RSID** | **Chr** | **Pos (hg19)** | **Effect Allele** | **Other Allele** | **Log OR (EUR)** | **P value (EUR)** | **Log OR (EAS)** | **P value (EAS)** | **Log OR (EAS/EUR)** | **P-value (EAS/EUR)** | **Initial publication** | **PMID** |
| --- | --- | --- | --- | --- | --- | --- | --- | --- | --- | --- | --- | --- |
| rs61776719 | 1 | 38461319 | C | A | 0.068 | 2.19E-10 | 0.049 | 6.78E-04 | 0.060 | 5.25E-11 | Law et al. 2019 NC | 31089142 |
| rs12144319 | 1 | 55246035 | C | T | 0.068 | 5.50E-06 | 0.022 | 1.23E-01 | 0.044 | 1.01E-05 | Huyghe et al. 2019 Nature Genetics | 30510241 |
| rs12143541 | 1 | 55247852 | G | A | 0.095 | 9.44E-10 | 0.078 | 8.00E-02 | 0.093 | 1.14E-09 | Law et al. 2019 NC | 31089142 |
| rs7542665 | 1 | 62673037 | C | T | 0.010 | 2.30E-01 | 0.080 | 5.20E-08 | 0.032 | 9.69E-05 | Lu et al. 2018 Gastroenterology | 30529582 |
| rs6678517 | 1 | 183002639 | A | G | 0.095 | 5.30E-14 | 0.057 | 4.13E-05 | 0.076 | 9.61E-15 | Huyghe et al. 2019 Nature Genetics | 30510241 |
| rs17011141 | 1 | 222112634 | G | A | 0.086 | 3.20E-08 | 0.026 | 2.26E-01 | 0.068 | 7.28E-09 | Huyghe et al. 2019 Nature Genetics | 30510241 |
| rs7606562 | 2 | 48686695 | T | A | 0.010 | 3.50E-01 | 0.093 | 1.27E-07 | 0.031 | 4.63E-04 | Lu et al. 2018 Gastroenterology | 30529582 |
| rs448513 | 2 | 159964552 | C | T | 0.049 | 5.80E-04 | 0.053 | 2.49E-04 | 0.051 | 6.90E-07 | Huyghe et al. 2019 Nature Genetics | 30510241 |
| rs11893063 | 2 | 199601925 | A | G | 0.068 | 9.34E-09 | 0.052 | 2.41E-04 | 0.061 | 2.41E-11 | Law et al. 2019 NC | 31089142 |
| rs983402 | 2 | 199781586 | T | C | 0.077 | 1.00E-08 | 0.044 | 2.95E-02 | 0.066 | 1.21E-08 | Huyghe et al. 2019 Nature Genetics | 30510241 |
| rs7593422 | 2 | 200131695 | T | A | 0.068 | 3.56E-11 | 0.019 | 2.13E-01 | 0.049 | 1.51E-07 | Law et al. 2019 NC | 31089142 |
| rs992157 | 2 | 219154781 | A | G | 0.095 | 3.15E-08 | 0.034 | 1.40E-02 | 0.059 | 1.77E-08 | Orlando et al. 2016 | 27005424 |
| rs35470271 | 3 | 40915239 | G | A | 0.068 | 9.50E-05 | 0.054 | 6.04E-04 | 0.059 | 9.38E-07 | Huyghe et al. 2019 Nature Genetics | 30510241 |
| rs9831861 | 3 | 53088285 | G | T | 0.068 | 4.17E-10 | 0.027 | 5.27E-02 | 0.054 | 4.29E-12 | Law et al. 2019 NC | 31089142 |
| rs6781752 | 3 | 66365163 | A | G | 0.049 | 1.90E-03 | 0.013 | 5.60E-01 | 0.038 | 1.73E-03 | Huyghe et al. 2019 Nature Genetics | 30510241 |
| rs12635946 | 3 | 112916918 | C | T | 0.077 | 1.02E-11 | 0.056 | 2.42E-04 | 0.071 | 8.02E-19 | Law et al. 2019 NC | 31089142 |
| rs72942485 | 3 | 112999560 | G | A | 0.207 | 1.50E-05 | 0.102 | 3.38E-04 | 0.130 | 1.14E-07 | Huyghe et al. 2019 Nature Genetics | 30510241 |
| rs10049390 | 3 | 133701119 | A | G | 0.068 | 1.80E-05 | 0.061 | 8.37E-06 | 0.064 | 8.20E-11 | Huyghe et al. 2019 Nature Genetics | 30510241 |
| rs113569514 | 3 | 133748789 | T | C | 0.058 | 1.60E-05 | 0.095 | 3.24E-11 | 0.073 | 1.42E-15 | Lu et al. 2018 Gastroenterology | 30529582 |
| rs10936599 | 3 | 169492101 | C | T | 0.077 | 3.39E-08 | 0.057 | 1.83E-05 | 0.065 | 4.72E-10 | Houlston et al.2010 | 20972440 |
| rs13149359 | 4 | 94938618 | A | C | 0.049 | 2.30E-04 | 0.011 | 5.02E-01 | 0.032 | 3.46E-03 | Huyghe et al. 2019 Nature Genetics | 30510241 |
| rs17035289 | 4 | 106048291 | T | C | 0.095 | 2.73E-10 | 0.045 | 9.42E-03 | 0.075 | 3.45E-12 | Law et al. 2019 NC | 31089142 |
| rs35509282 | 4 | 163333405 | A | T | 0.425 | 8.20E-09 | 0.021 | 1.23E-01 | 0.036 | 7.41E-03 | Schmit et al. 2014 | 25023989 |
| rs2735940 | 5 | 1296486 | G | A | 0.086 | 3.60E-11 | 0.060 | 5.01E-05 | 0.074 | 4.43E-13 | Huyghe et al. 2019 Nature Genetics | 30510241 |
| rs7708610 | 5 | 40102443 | A | G | 0.058 | 5.56E-06 | 0.044 | 6.80E-03 | 0.053 | 3.34E-08 | Huyghe et al. 2019 Nature Genetics | 30510241 |
| rs12514517 | 5 | 40280076 | A | G | 0.095 | 1.40E-12 | 0.125 | 1.26E-04 | 0.101 | 2.87E-12 | Huyghe et al. 2019 Nature Genetics | 30510241 |
| rs58791712 | 5 | 40281797 | GT | G | 0.095 | 7.30E-14 | 0.100 | 3.80E-02 | 0.096 | 3.20E-17 | Schmit et al. 2018 | 29917119 |
| rs12659017 | 5 | 125988175 | G | A | -0.010 | 4.60E-01 | 0.090 | 1.53E-08 | 0.019 | 2.51E-02 | Lu et al. 2018 Gastroenterology | 30529582 |
| rs639933 | 5 | 134467751 | C | A | 0.068 | 1.14E-09 | 0.117 | 8.51E-18 | 0.089 | 2.72E-23 | Law et al. 2019 NC | 31089142 |
| rs2070699 | 6 | 12292772 | T | G | 0.068 | 3.88E-09 | 0.047 | 1.06E-03 | 0.059 | 1.21E-10 | Law et al. 2019 NC | 31089142 |
| rs1476570 | 6 | 29809860 | A | G | 0.010 | 4.40E-01 | 0.115 | 2.14E-09 | 0.033 | 2.56E-04 | Lu et al. 2018 Gastroenterology | 30529582 |
| rs2516420 | 6 | 31449620 | C | T | 0.166 | 4.76E-11 | 0.099 | 4.99E-02 | 0.152 | 5.15E-11 | Huyghe et al. 2019 Nature Genetics | 30510241 |
| rs3830041 | 6 | 32191339 | T | C | 0.000 | 9.20E-01 | 0.147 | 1.36E-08 | 0.038 | 3.73E-03 | Lu et al. 2018 Gastroenterology | 30529582 |
| rs9271695 | 6 | 32593080 | G | A | 0.086 | 1.70E-07 | 0.048 | 1.39E-02 | 0.071 | 2.25E-08 | Huyghe et al. 2019 Nature Genetics | 30510241 |
| rs6906359 | 6 | 35528378 | C | T | 0.104 | 3.40E-08 | 0.031 | 2.84E-01 | 0.075 | 2.60E-05 | Schmit et al. 2018 | 29917119 |
| rs16878812 | 6 | 35569562 | A | G | 0.068 | 1.20E-03 | 0.155 | 4.52E-02 | 0.073 | 8.77E-05 | Huyghe et al. 2019 Nature Genetics | 30510241 |
| rs9470361 | 6 | 36623379 | A | G | 0.068 | 1.30E-05 | 0.082 | 7.48E-06 | 0.073 | 9.07E-11 | Huyghe et al. 2019 Nature Genetics | 30510241 |
| rs4711689 | 6 | 41692812 | A | G | 0.010 | 6.90E-01 | 0.082 | 9.11E-06 | 0.045 | 5.04E-04 | Zeng C, Gastroenterology, 2016 | 26965516 |
| rs62404968 | 6 | 55714314 | C | T | 0.086 | 8.60E-10 | 0.079 | 2.10E-02 | 0.085 | 1.08E-08 | Schmit et al. 2018 | 29917119 |
| rs6928864 | 6 | 105966894 | C | A | 0.122 | 1.37E-08 | 0.061 | 4.35E-02 | 0.100 | 2.35E-08 | Law et al. 2019 NC | 31089142 |
| rs12672022 | 7 | 45136423 | T | C | 0.058 | 4.40E-04 | 0.050 | 7.39E-03 | 0.055 | 1.22E-05 | Huyghe et al. 2019 Nature Genetics | 30510241 |
| rs4308634 | 7 | 46887213 | G | C | 0.039 | 3.54E-05 | 0.052 | 2.56E-04 | 0.042 | 1.62E-10 | Lu et al. 2019 | 31826910 |
| rs3801081 | 7 | 47511161 | G | A | 0.077 | 2.00E-11 | 0.062 | 6.17E-04 | 0.072 | 1.87E-13 | Law et al. 2019 NC | 31089142 |
| rs60911071 | 8 | 23664632 | G | C | 0.049 | 3.20E-02 | 0.073 | 1.55E-07 | 0.066 | 1.67E-08 | Lu et al. 2019 | 31826910 |
| rs3133285 | 8 | 117629411 | G | C | 0.068 | 3.40E-05 | 0.106 | 1.21E-14 | 0.090 | 1.31E-17 | Huyghe et al. 2019 Nature Genetics | 30510241 |
| rs6983267 | 8 | 128413305 | G | T | 0.157 | 9.50E-36 | 0.155 | 1.19E-31 | 0.156 | 4.67E-63 | Huyghe et al. 2019 Nature Genetics | 30510241 |
| rs4313119 | 8 | 128571855 | G | T | 0.039 | 4.36E-03 | 0.041 | 8.52E-03 | 0.040 | 1.80E-04 | Huyghe et al. 2019 Nature Genetics | 30510241 |
| rs1412834 | 9 | 22110131 | T | C | 0.077 | 4.13E-14 | 0.034 | 1.26E-02 | 0.058 | 4.85E-11 | Law et al. 2019 NC | 31089142 |
| rs62558833 | 9 | 34039002 | T | C | 0.030 | 1.70E-03 | 0.058 | 9.06E-05 | 0.038 | 3.23E-06 | Lu et al. 2019 | 31826910 |
| rs34405347 | 9 | 101679752 | T | G | 0.086 | 1.50E-04 | 0.052 | 1.98E-03 | 0.065 | 7.37E-07 | Huyghe et al. 2019 Nature Genetics | 30510241 |
| rs10980628 | 9 | 113671403 | C | T | 0.077 | 1.30E-06 | 0.032 | 4.86E-02 | 0.057 | 7.91E-08 | Huyghe et al. 2019 Nature Genetics | 30510241 |
| rs11255841 | 10 | 8739580 | T | A | 0.095 | 2.40E-12 | 0.155 | 2.17E-27 | 0.124 | 1.27E-35 | Huyghe et al. 2019 Nature Genetics | 30510241 |
| rs10821907 | 10 | 52648454 | C | T | 0.068 | 9.90E-05 | 0.031 | 3.42E-01 | 0.058 | 4.10E-04 | Huyghe et al. 2019 Nature Genetics | 30510241 |
| rs704017 | 10 | 80819132 | G | A | 0.077 | 1.50E-09 | 0.087 | 4.33E-10 | 0.082 | 1.66E-16 | Huyghe et al. 2019 Nature Genetics | 30510241 |
| rs6584283 | 10 | 101290301 | C | T | 0.030 | 3.60E-04 | 0.089 | 2.02E-11 | 0.051 | 1.41E-10 | Lu et al. 2018 Gastroenterology | 30529582 |
| rs1035209 | 10 | 101345366 | T | C | 0.113 | 4.45E-11 | 0.071 | 4.09E-05 | 0.091 | 3.85E-13 | Whiffin et al. 2014 | 24737748 |
| rs4919687 | 10 | 104595248 | G | A | 0.049 | 2.00E-03 | 0.069 | 1.89E-04 | 0.058 | 3.28E-06 | Zeng C, Gastroenterology, 2016 | 26965516 |
| rs12241008 | 10 | 114280702 | C | T | 0.086 | 1.50E-03 | 0.102 | 1.63E-11 | 0.098 | 1.47E-13 | Wang H, Nat Comm, 2014 | 25105248 |
| rs11196172 | 10 | 114726843 | A | G | 0.058 | 1.10E-01 | 0.124 | 2.12E-16 | 0.113 | 2.27E-16 | Zhang B, Nat Genet, 2014 | 24836286 |
| rs174537 | 11 | 61552680 | G | T | 0.068 | 7.39E-05 | 0.097 | 3.70E-12 | 0.085 | 1.96E-15 | Zhang B, Nat Genet, 2014 | 24836286 |
| rs3824999 | 11 | 74345550 | G | T | 0.077 | 3.65E-10 | 0.066 | 1.96E-06 | 0.072 | 1.09E-15 | Dunlop MG, et al. 2012 | 22634755 |
| rs2186607 | 11 | 101656397 | T | A | 0.049 | 3.30E-05 | 0.003 | 8.98E-01 | 0.036 | 4.71E-04 | Huyghe et al. 2019 Nature Genetics | 30510241 |
| rs3802842 | 11 | 111171709 | C | A | 0.104 | 5.82E-10 | 0.063 | 3.11E-06 | 0.080 | 8.84E-15 | Tenesa et al. 2008 | 18372901 |
| rs35808169 | 12 | 4368607 | C | T | 0.058 | 1.00E-03 | 0.115 | 3.84E-13 | 0.088 | 2.00E-14 | Huyghe et al. 2019 Nature Genetics | 30510241 |
| rs3217810 | 12 | 4388271 | T | C | 0.113 | 4.00E-08 | 0.067 | 3.77E-01 | 0.110 | 3.05E-08 | Huyghe et al. 2019 Nature Genetics | 30510241 |
| rs3217874 | 12 | 4400808 | T | C | 0.068 | 1.98E-06 | 0.058 | 2.23E-05 | 0.064 | 1.93E-12 | Huyghe et al. 2019 Nature Genetics | 30510241 |
| rs10849432 | 12 | 6385727 | T | C | 0.030 | 5.00E-01 | 0.076 | 1.93E-05 | 0.068 | 2.72E-05 | Zhang B, Nat Genet, 2014 | 24836286 |
| rs10849438 | 12 | 6412036 | G | T | 0.113 | 1.04E-10 | 0.028 | 6.77E-02 | 0.062 | 7.95E-08 | Law et al. 2019 NC | 31089142 |
| rs2250430 | 12 | 6421174 | T | A | 0.058 | 7.50E-05 | 0.035 | 5.18E-02 | 0.049 | 1.26E-05 | Huyghe et al. 2019 Nature Genetics | 30510241 |
| rs11064437 | 12 | 6982162 | C | T | 0.157 | 3.30E-01 | 0.039 | 8.91E-03 | 0.040 | 7.29E-03 | Zeng C, Gastroenterology, 2016 | 26965516 |
| rs2238126 | 12 | 12009741 | G | A | 0.174 | 3.40E-02 | 0.035 | 1.11E-02 | 0.035 | 9.43E-03 | Wang M, Nat Comm, 2016 | 27145994 |
| rs2730985 | 12 | 43130624 | G | A | 0.049 | 5.50E-09 | 0.079 | 1.30E-08 | 0.059 | 1.80E-13 | Lu et al. 2018 Gastroenterology | 30529582 |
| rs12372718 | 12 | 51171090 | G | A | 0.095 | 2.90E-12 | 0.048 | 6.87E-04 | 0.076 | 2.30E-17 | Huyghe et al. 2019 Nature Genetics | 30510241 |
| rs7398375 | 12 | 57540848 | C | G | 0.086 | 3.91E-10 | 0.071 | 1.12E-05 | 0.078 | 8.13E-12 | Law et al. 2019 NC | 31089142 |
| rs11108175 | 12 | 96050887 | A | G | 0.049 | 2.60E-07 | 0.075 | 8.54E-06 | 0.055 | 5.13E-11 | Lu et al. 2019 | 31826910 |
| rs9634162 | 12 | 115098094 | A | G | 0.039 | 7.50E-07 | 0.061 | 2.03E-05 | 0.044 | 2.04E-11 | Lu et al. 2019 | 31826910 |
| rs72013726 | 12 | 115890835 | C | CACA | 0.077 | 5.00E-11 | 0.047 | 1.31E-02 | 0.064 | 3.09E-07 | Schmit et al. 2018 | 29917119 |
| rs7300312 | 12 | 115890922 | C | T | 0.068 | 9.20E-07 | 0.037 | 6.80E-03 | 0.055 | 1.52E-09 | Huyghe et al. 2019 Nature Genetics | 30510241 |
| rs55990915 | 12 | 117763309 | A | C | 0.039 | 6.60E-02 | 0.051 | 9.75E-02 | 0.042 | 5.24E-03 | Huyghe et al. 2019 Nature Genetics | 30510241 |
| rs10161980 | 13 | 34093518 | C | G | 0.077 | 4.70E-09 | 0.030 | 4.32E-02 | 0.054 | 1.02E-07 | Schmit et al. 2018 | 29917119 |
| rs7333607 | 13 | 37462010 | G | A | 0.068 | 4.40E-06 | 0.070 | 1.47E-03 | 0.068 | 1.19E-08 | Huyghe et al. 2019 Nature Genetics | 30510241 |
| rs45597035 | 13 | 73649152 | A | G | 0.077 | 2.16E-10 | 0.015 | 3.80E-01 | 0.057 | 5.33E-09 | Law et al. 2019 NC | 31089142 |
| rs1886450 | 13 | 73986628 | G | A | 0.020 | 9.70E-03 | 0.097 | 2.71E-13 | 0.038 | 3.31E-09 | Lu et al. 2018 Gastroenterology | 30529582 |
| rs1330889 | 13 | 78609615 | C | T | 0.104 | 6.50E-10 | 0.055 | 2.73E-02 | 0.090 | 3.59E-11 | Law et al. 2019 NC | 31089142 |
| rs35107139 | 14 | 54419106 | C | A | 0.086 | 9.90E-12 | 0.077 | 4.39E-07 | 0.083 | 3.74E-19 | Huyghe et al. 2019 Nature Genetics | 30510241 |
| rs16969681 | 15 | 32993111 | T | C | 0.157 | 1.10E-12 | 0.077 | 1.65E-08 | 0.099 | 7.61E-18 | Huyghe et al. 2019 Nature Genetics | 30510241 |
| rs17816465 | 15 | 33156386 | A | G | 0.095 | 4.02E-10 | 0.170 | 1.21E-04 | 0.104 | 6.92E-12 | Huyghe et al. 2019 Nature Genetics | 30510241 |
| rs4776316 | 15 | 67007813 | A | G | 0.077 | 1.11E-08 | 0.022 | 2.32E-01 | 0.061 | 1.04E-09 | Law et al. 2019 NC | 31089142 |
| rs56324967 | 15 | 67402824 | C | T | 0.077 | 9.80E-08 | 0.039 | 1.27E-02 | 0.060 | 1.16E-08 | Huyghe et al. 2019 Nature Genetics | 30510241 |
| rs10152518 | 15 | 68177162 | G | A | 0.077 | 3.24E-08 | 0.026 | 7.02E-02 | 0.052 | 2.81E-07 | Law et al. 2019 NC | 31089142 |
| rs9924886 | 16 | 68743939 | A | C | 0.049 | 2.40E-04 | 0.059 | 6.84E-04 | 0.053 | 1.86E-06 | Huyghe et al. 2019 Nature Genetics | 30510241 |
| rs4341754 | 16 | 80039621 | G | C | 0.049 | 5.20E-08 | 0.088 | 1.89E-10 | 0.062 | 8.89E-15 | Lu et al. 2018 Gastroenterology | 30529582 |
| rs2696839 | 16 | 86340448 | G | C | 0.058 | 2.00E-08 | 0.035 | 2.36E-02 | 0.050 | 1.65E-07 | Schmit et al. 2018 | 29917119 |
| rs62042090 | 16 | 86703949 | T | C | 0.058 | 2.40E-04 | 0.051 | 2.12E-03 | 0.055 | 4.19E-07 | Huyghe et al. 2019 Nature Genetics | 30510241 |
| rs4968127 | 17 | 809643 | G | A | 0.068 | 4.00E-07 | 0.070 | 3.03E-07 | 0.069 | 3.14E-12 | Huyghe et al. 2019 Nature Genetics | 30510241 |
| rs1078643 | 17 | 10707241 | A | G | 0.086 | 1.10E-07 | 0.117 | 1.61E-11 | 0.101 | 3.12E-17 | Huyghe et al. 2019 Nature Genetics | 30510241 |
| rs983318 | 17 | 70413253 | A | G | 0.049 | 8.00E-04 | 0.027 | 4.25E-01 | 0.046 | 6.95E-04 | Huyghe et al. 2019 Nature Genetics | 30510241 |
| rs75954926 | 17 | 81061048 | G | A | 0.086 | 4.80E-09 | 0.081 | 1.37E-03 | 0.085 | 4.51E-12 | Huyghe et al. 2019 Nature Genetics | 30510241 |
| rs11874392 | 18 | 46453156 | A | T | 0.174 | 2.10E-41 | 0.194 | 7.01E-43 | 0.183 | 3.51E-82 | Huyghe et al. 2019 Nature Genetics | 30510241 |
| rs28840750 | 19 | 33519927 | T | G | 0.191 | 2.60E-10 | 0.187 | 1.46E-11 | 0.188 | 1.39E-19 | Huyghe et al. 2019 Nature Genetics | 30510241 |
| rs1800469 | 19 | 41860296 | G | A | 0.030 | 9.00E-02 | 0.052 | 8.32E-05 | 0.044 | 3.05E-05 | Zhang B, Nat Genet, 2014 | 24836286 |
| rs12979278 | 19 | 49218602 | T | C | 0.068 | 6.11E-10 | 0.016 | 6.83E-01 | 0.065 | 2.97E-12 | Law et al. 2019 NC | 31089142 |
| rs73068325 | 19 | 59079096 | T | C | 0.068 | 5.00E-05 | 0.059 | 5.75E-02 | 0.066 | 7.28E-06 | Huyghe et al. 2019 Nature Genetics | 30510241 |
| rs961253 | 20 | 6404281 | A | C | 0.113 | 2.00E-10 | 0.074 | 5.97E-04 | 0.097 | 3.22E-12 | Houlston et al. 2008 | 19011631 |
| rs994308 | 20 | 6603622 | C | T | 0.077 | 7.74E-09 | 0.029 | 9.97E-02 | 0.059 | 1.32E-07 | Huyghe et al. 2019 Nature Genetics | 30510241 |
| rs4813802 | 20 | 6699595 | G | T | 0.068 | 4.60E-07 | 0.099 | 1.81E-09 | 0.081 | 6.41E-14 | Huyghe et al. 2019 Nature Genetics | 30510241 |
| rs28488 | 20 | 6762221 | T | C | 0.068 | 1.78E-06 | 0.032 | 5.29E-02 | 0.053 | 1.24E-06 | Huyghe et al. 2019 Nature Genetics | 30510241 |
| rs2423279 | 20 | 7812350 | C | T | 0.068 | 1.00E-03 | 0.099 | 1.52E-11 | 0.089 | 1.89E-13 | Jia WH, Nat Genet, 2013 | 23263487 |
| rs2295444 | 20 | 33173883 | C | T | 0.077 | 3.30E-09 | 0.034 | 1.82E-02 | 0.060 | 7.10E-11 | Schmit et al. 2018 | 29917119 |
| rs2179593 | 20 | 42660286 | A | C | 0.068 | 4.62E-09 | 0.017 | 2.99E-01 | 0.050 | 1.73E-07 | Law et al. 2019 NC | 31089142 |
| rs6066825 | 20 | 47340117 | A | G | 0.077 | 1.70E-08 | 0.097 | 9.85E-11 | 0.086 | 5.17E-17 | Huyghe et al. 2019 Nature Genetics | 30510241 |
| rs1810502 | 20 | 49057488 | C | T | 0.077 | 1.02E-08 | 0.060 | 1.15E-05 | 0.070 | 8.20E-15 | Schmit et al. 2018 | 29917119 |
| rs13831 | 20 | 57475191 | G | A | 0.010 | 3.20E-01 | 0.079 | 2.66E-08 | 0.033 | 5.53E-05 | Lu et al. 2018 Gastroenterology | 30529582 |
| rs6061231 | 20 | 60956917 | C | A | 0.068 | 8.90E-05 | 0.166 | 1.36E-12 | 0.101 | 1.01E-13 | Zeng C, Gastroenterology, 2016 | 26965516 |
| rs2738783 | 20 | 62308612 | T | G | 0.049 | 3.30E-03 | 0.031 | 8.16E-02 | 0.042 | 2.22E-04 | Huyghe et al. 2019 Nature Genetics | 30510241 |

**eTable 3.** Associations of polygenic risk scores with colorectal cancer risk in the validation sets and the age-stratified China validation set.

| **PRS development methods** | **Korea validation set** | | **China validation set (All)** | | **China validation set (Younger, Age < 70.3)** | | **China validation set (Older, Age ≥ 70.3)** | |
| --- | --- | --- | --- | --- | --- | --- | --- | --- |
|  | **OR (95% CI)^1^** | **AUC (95% CI)** | **OR (95% CI)^1^** | **AUC (95% CI)** | **OR (95% CI)^1^** | **AUC (95% CI)** | **OR (95% CI)^1^** | **AUC (95% CI)** |
| **GWAS-reported index SNPs^2^** |  |  |  |  |  |  |  |  |
| **PRS_115-EAS_** | 1.63 (1.46 - 1.83) | 0.63 (0.60 - 0.66) | 1.51 (1.37 - 1.67) | 0.61 (0.59 - 0.64) | 1.54 (1.34 - 1.79) | 0.62 (0.58 - 0.66) | 1.47 (1.28 - 1.69) | 0.61 (0.57 - 0.64) |
| **PRS_115-EUR_** | 1.44 (1.29 - 1.60) | 0.60 (0.57 - 0.63) | 1.39 (1.26 - 1.53) | 0.59 (0.56 - 0.61) | 1.51 (1.31 - 1.75) | 0.61 (0.57 - 0.65) | 1.29 (1.13 - 1.48) | 0.57 (0.53 - 0.61) |
| **PRS_115-EAS/EUR_** | 1.68 (1.50 - 1.89) | 0.64 (0.61 - 0.67) | 1.50 (1.36 - 1.66) | 0.61 (0.58 - 0.64) | 1.62 (1.40 - 1.88) | 0.63 (0.59 - 0.67) | 1.39 (1.21 - 1.60) | 0.59 (0.56 - 0.63) |
| **SNPs selected by fine-mapping^3^** |  |  |  |  |  |  |  |  |
| **PRS_168_ (P < 1.0 x 10^-3^)** | 1.54 (1.38 - 1.72) | 0.62 (0.59 - 0.65) | 1.31 (1.19 - 1.45) | 0.58 (0.55 - 0.61) | 1.23 (1.07 - 1.41) | 0.56 (0.53 - 0.60) | 1.39 (1.21 - 1.59) | 0.59 (0.55 - 0.63) |
| **PRS_129_ (P < 1.0 x 10^-4^)** | 1.37 (1.23 - 1.52) | 0.59 (0.56 - 0.62) | 1.31 (1.19 - 1.44) | 0.58 (0.55 - 0.60) | 1.19 (1.04 - 1.37) | 0.56 (0.52 - 0.60) | 1.38 (1.20 - 1.58) | 0.59 (0.55 - 0.62) |
| **PRS_115_ (P < 1.0 x 10^-5^)** | 1.38 (1.24 - 1.54) | 0.59 (0.56 - 0.62) | 1.27 (1.15 - 1.40) | 0.57 (0.54 - 0.59) | 1.20 (1.05 - 1.38) | 0.56 (0.52 - 0.60) | 1.30 (1.14 - 1.49) | 0.57 (0.53 - 0.61) |
| **Genome-wide risk prediction algorithm^4^** |  |  |  |  |  |  |  |  |
| **PRS_LDpred2-Inf_** | 1.23 (1.10 - 1.36) | 0.56 (0.53 - 0.59) | 1.22 (1.11 - 1.34) | 0.56 (0.53 - 0.58) | 1.27 (1.11 - 1.46) | 0.57 (0.53 - 0.60) | 1.18 (1.03 - 1.34) | 0.54 (0.51 - 0.58) |
| **PRS_LDpred2-grid-nosp_** | 1.34 (1.21 - 1.50) | 0.59 (0.56 - 0.62) | 1.28 (1.16 - 1.41) | 0.57 (0.54 - 0.59) | 1.37 (1.20 - 1.58) | 0.59 (0.55 - 0.62) | 1.25 (1.09 - 1.43) | 0.56 (0.52 - 0.60) |
| **PRS_LDpred2-grid-sp_** | 1.43 (1.29 - 1.60) | 0.60 (0.57 - 0.63) | 1.32 (1.20 - 1.45) | 0.58 (0.55 - 0.60) | 1.35 (1.18 - 1.56) | 0.58 (0.54 - 0.62) | 1.23 (1.08 - 1.41) | 0.56 (0.52 - 0.60) |
| **PRS_LDpred2-Auto_** | 1.31 (1.18 - 1.46) | 0.58 (0.55 - 0.61) | 1.27 (1.15 - 1.40) | 0.57 (0.54 - 0.59) | 1.34 (1.16 - 1.54) | 0.58 (0.54 - 0.62) | 1.21 (1.06 - 1.38) | 0.55 (0.51 - 0.59) |
| **PRS_PRS-CS_** | 1.34 (1.20 - 1.49) | 0.58 (0.55 - 0.61) | 1.35 (1.23 - 1.49) | 0.58 (0.55 - 0.61) | 1.44 (1.25 - 1.66) | 0.60 (0.56 - 0.64) | 1.31 (1.15 - 1.51) | 0.58 (0.54 - 0.61) |

**eTable 4.** 10-Year Absolute Risk (%) for individuals at age 50 years with different PRS_115-EAS/EUR_ cores in China, South Korea, and Japan.

| **PRS Percentile** | **China** | **South Korea** | **Japan** |
| --- | --- | --- | --- |
| **>99** | 1.50 | 1.65 | 2.56 |
| **(95, 99]** | 1.10 | 1.20 | 1.87 |
| **(40, 60]** | 0.47 | 0.52 | 0.80 |
| **(1, 5]** | 0.22 | 0.25 | 0.38 |
| **≤1** | 0.16 | 0.18 | 0.27 |
